# Amyotrophic lateral sclerosis progression is tracked in plasma by systemic proteins independent of motor neurons

**DOI:** 10.64898/2026.11.15.26368153

**Authors:** Caitlin A. Finney, Artur Shvetcov

## Abstract

Blood biomarkers that change as patients with amyotrophic lateral sclerosis (ALS) decline would allow progression to be monitored more objectively. Plasma proteomic studies have identified proteins that differ between patients and controls, but which plasma proteins change as patients decline, and whether they come from motor neurons, is unknown. Here, we leveraged the Answer ALS cohort, with serial SomaScan plasma proteomics, repeated ALS Functional Rating Scale-Revised assessments, and induced pluripotent stem cell-derived motor neurons (iMNs) from the same patients. At baseline, 77 plasma proteins differed between 245 patients and 76 controls. These were dominated by muscle proteins, including PDLIM3, ACTN2, and MYOM2, and reflected disease severity. Using a two-stage longitudinal design, we identified 72 progression markers whose within-patient change tracked the rate of functional decline, led by CLEC3B, ANTXR2, and ART3. These were largely distinct from the most elevated muscle proteins and were secreted or shed proteins spread across many tissues. Both sets replicated in the independent Target ALS cohort. A motor neuron expression map built from the iMNs showed that motor neuron proteins were underrepresented in both sets and that expression in iMNs did not predict tracking of decline. Baseline serum neurofilament light chain (NfL) predicted how quickly patients declined but its change did not track decline, whereas change in the progression markers did so within every stratum of baseline NfL. Neither tracked decline in the fastest progressing patients. This positions baseline NfL for prognosis and stratification and systemic plasma proteins for monitoring.

## Introduction

Amyotrophic lateral sclerosis (ALS) is a fatal neurodegenerative disease of upper and lower motor neurons in which the rate of functional decline varies several fold between patients (*1*). This heterogeneity is one of the main obstacles to clinical trials. Functional decline is measured using the ALS Functional Rating Scale-Revised (ALSFRS-R) (*2*), which is the primary endpoint in most trials. However, the ALSFRS-R changes slowly relative to the length of a trial, is not linear over the course of disease, and combines bulbar, motor, and respiratory function into a single score (*3–5*). Blood biomarkers that change as patients decline would allow progression to be monitored more objectively.

Neurofilament light chain (NfL) is the most established blood biomarker in ALS (*6, 7*). Serum and plasma NfL are elevated in patients with ALS, rise before symptom onset in genetic carriers, and predict how quickly patients will decline and how long they will survive (*8–12*). A reduction in plasma NfL was also central to the approval of tofersen for SOD1 ALS, making NfL the first blood biomarker used as a surrogate endpoint in the disease (*13, 14*). Despite this, NfL levels are relatively stable over time within individual patients (*9, 10*), and whether within-patient change in NfL tracks the rate of functional decline as it occurs has not been established.

Large-scale plasma proteomic studies have identified hundreds of proteins that differ between patients with ALS and controls. Across SomaScan and Olink platforms, these differences are dominated by skeletal muscle proteins and include proteins that change years before symptom onset (*15–17*), and metabolic and immune changes in the periphery are associated with faster progression (*18, 19*). In these studies, however, the tissue origin of plasma proteins has been inferred from tissue expression databases, which do not include motor neurons, and progression has largely been studied as the association of a baseline protein level with subsequent decline or survival (*16, 20*). Proteomic differences between fast and slow progressors have also been reported in longitudinal cerebrospinal fluid (CSF) (*21*), suggesting that the biology of decline may differ with rate of progression. It is unknown whether the plasma proteins that change as ALS progresses come from motor neurons, whether they track functional decline, and whether this holds across individual differences in progression rates.

To address this, we leveraged the Answer ALS cohort (*22*). Here, the plasma proteome was measured by SomaScan in serial samples alongside repeated ALSFRS-R assessments, serum NfL was measured by immunoassay, and induced pluripotent stem cell (iPSC)-derived motor neurons (iMNs) from the same patients were profiled by RNA-sequencing, mass spectrometry, and ATAC-sequencing (*22, 23*). Using a two-stage longitudinal design, we related each patient’s rate of change in each plasma protein to their rate of functional decline and built a motor neuron expression map of the plasma proteome from the iMN data. We replicated these findings on mass spectrometry and Olink in the independent Target ALS Natural History Study (*24*). Lastly, we compared change in the progression markers with baseline and change in serum NfL and examined how both tracked decline across patients who progressed at different rates (Fig. 1A).

**Figure 1.**
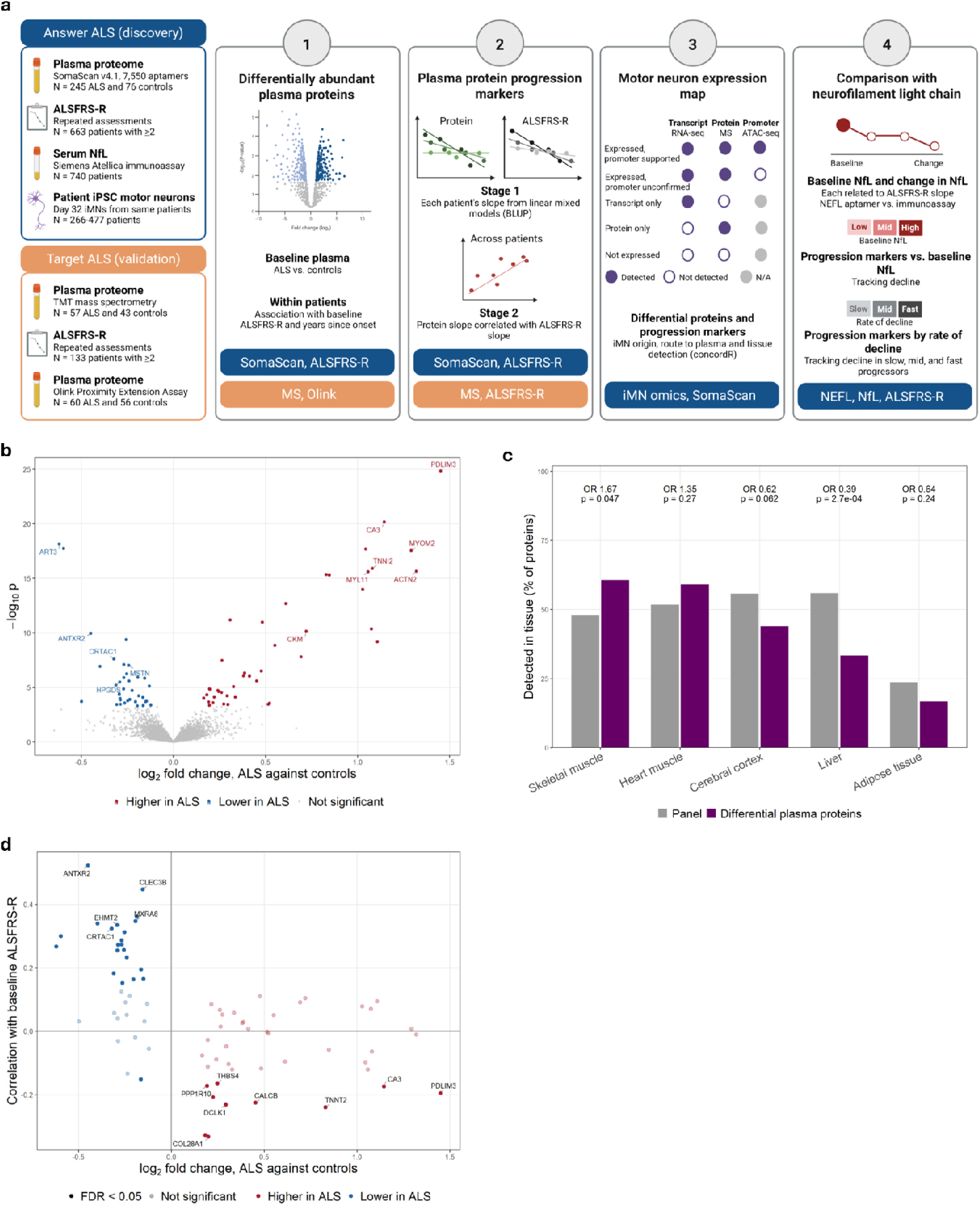
Differential plasma proteins in ALS are dominated by muscle proteins and reflect disease severity. (**A**) Study design, with Answer ALS as the discovery cohort and the Target ALS Natural History Study as the validation cohort. Differential plasma proteins were identified at baseline (1), progression markers were identified using a two-stage design (2), both were tested against a motor neuron expression map built from patient iMNs (3), and the progression markers were compared with serum NfL (4). (**B**) Volcano plot of baseline differences between patients with ALS (n = 245) and controls (n = 76) for the 7,550 SomaScan aptamers, from linear models adjusted for sex and age. Aptamers higher (red) or lower (blue) in ALS at a false discovery rate (FDR) of 5% are colored and selected proteins are labeled. (**C**) Percentage of the differential plasma proteins (purple) and of the panel (gray) detected in each tissue. Odds ratios (OR) and p-values compare the differential plasma proteins with the panel. (**D**) Pearson correlation of each of the 77 differential plasma proteins with baseline ALSFRS-R score in patients with ALS (n = 244), plotted against its log_2_ fold change between ALS and controls. Dark points indicate an FDR below 0.05 and light points were not significant. Abbreviations: ALSFRS-R: ALS Functional Rating Scale-Revised, BLUP: best linear unbiased predictor, MS: mass spectrometry.

## Results

### Plasma proteins that distinguish ALS from controls are dominated by muscle proteins and reflect disease severity

We first compared the baseline plasma proteomes of 245 patients with ALS and 76 controls from the Answer ALS cohort, adjusting for sex and age (Table S1). Of the 7,550 SomaScan aptamers measured in plasma, 77 differed at a false discovery rate (FDR) of 5%, with 42 higher and 35 lower in patients (Fig. 1B; Table S2). The largest elevations were in skeletal and cardiac muscle proteins, including PDZ and LIM domain protein 3 (PDLIM3, log_2_FC = 1.45), α-actinin-2 (ACTN2, log_2_FC = 1.32), myomesin-2 (MYOM2, log_2_FC = 1.29), carbonic anhydrase 3 (CA3, log_2_FC = 1.15), fast skeletal troponin I (TNNI2, log_2_FC = 1.08), myosin light chain 11 (MYL11, log_2_FC = 1.06), and muscle creatine kinase (CKM, log_2_FC = 0.72; Fig. 1B). Muscle proteins decreased in patients included myostatin (MSTN, log_2_FC = -0.24) and ecto-ADP-ribosyltransferase 3 (ART3, log_2_FC = -0.62), anthrax toxin receptor 2 (ANTXR2, log_2_FC = -0.45). Non-muscle decreased proteins were cartilage acidic protein 1 (CRTAC1, log_2_FC = -0.32) and hematopoietic prostaglandin D synthase (HPGDS, log_2_FC = -0.29). The differential plasma proteins were enriched for detection in skeletal muscle (61% versus 48% of the panel, OR = 1.67, *p* = 0.047) and depleted for detection in liver (33% versus 56%, OR = 0.39, *p* = 3 x 10^-4^; Fig. 1C).

To determine whether these differences reflected disease severity or duration, we related each differential plasma protein to baseline ALSFRS-R score and to years since symptom onset. In 244 patients, 32 of the 77 differential plasma proteins were associated with the score at an FDR of 5% (Fig. 1D; Table S3). The strongest associations were for proteins that were lower in patients and lowest in the most impaired, led by ANTXR2 (r = 0.52), tetranectin (CLEC3B, r = 0.45), and matrix remodeling-associated protein 8 (MXRA8, r = 0.35). Only one differential plasma protein, osteomodulin (OMD), was associated with years since symptom onset (r = 0.24, FDR = 0.014).

The differential plasma proteins are therefore dominated by muscle proteins, as reported in other cohorts (*15, 16*), and largely reflect how impaired a patient is at the time of sampling and not how long they have had the disease.

### Within-patient change in plasma proteins tracks the rate of functional decline in ALS

To identify plasma proteins that change with disease progression, we used a two-stage design. Here, each patient’s rate of functional decline was estimated from longitudinal ALSFRS-R scores (n = 663) and their rate of change in each plasma protein from serial plasma draws (n = 229) using linear mixed-effects models, and the two were related across patients. We chose this design over alternative methods because, in simulations calibrated to the cohort, it detected most true progression markers while controlling the FDR, whereas the alternatives either missed most markers or returned a high proportion of false positives (Fig. S1 to S3). Of the 7,550 aptamers, 729 were nominally associated with the rate of decline and 72 remained significant at an FDR of 5% (Fig. 2A; Table S4). The strongest associations were for CLEC3B (r = 0.56), ANTXR2 (r = 0.55), ART3 (r = 0.45), HPGDS (r = 0.43), and myocilin (MYOC, r = 0.43), and for each of these, faster functional decline was accompanied by a faster protein decrease in plasma. Of the 66 differential plasma proteins with a unique gene symbol, 23 (35%) were also progression markers. These included 16 (55%) of the 29 that were lower in patients, such as CLEC3B, ANTXR2, ART3, CRTAC1, and HPGDS, compared with seven (19%) of the 37 that were higher in patients (OR = 5.27, *p* = 0.004; Fig. 2B). The muscle proteins with the largest elevations, including PDLIM3, CA3, TNNI2, and MYL11, were not progression markers.

**Figure 2.**
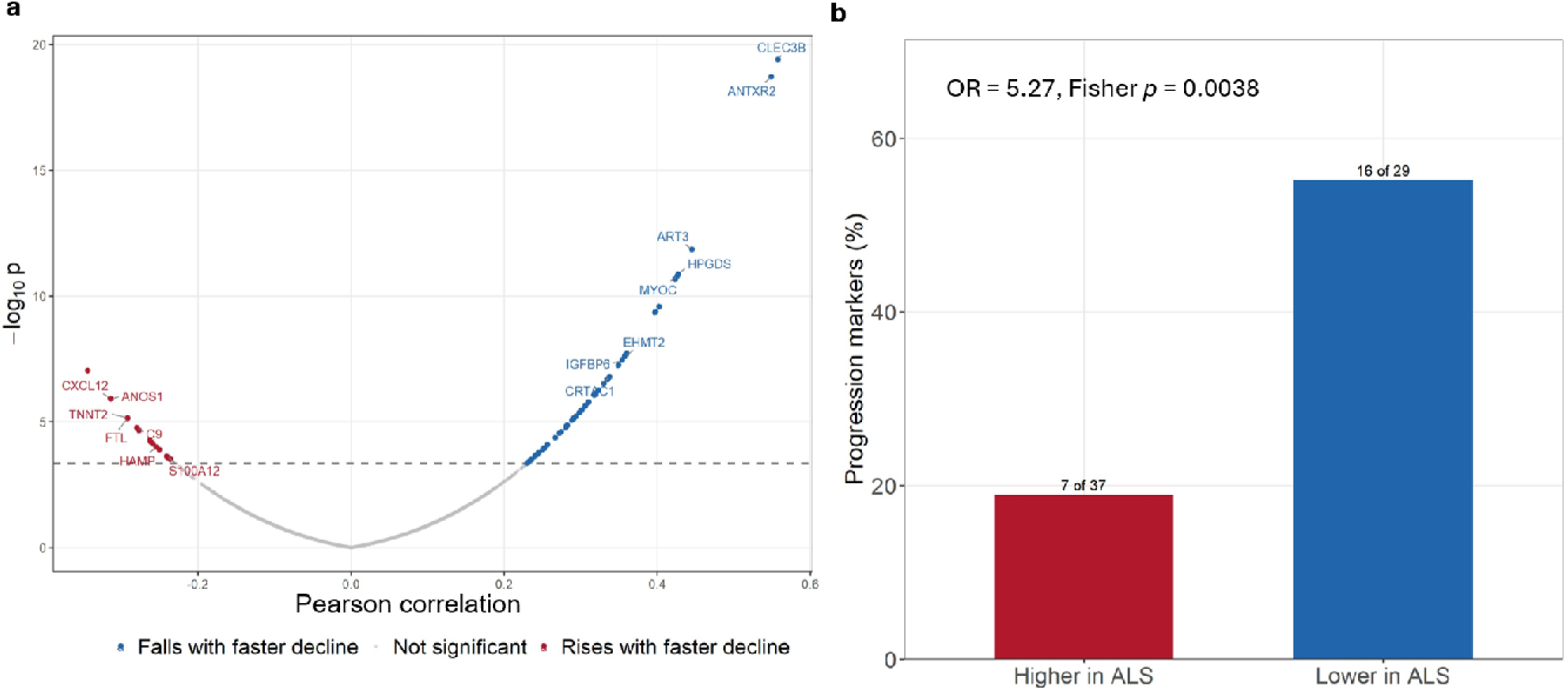
Within-patient change in plasma proteins tracks the rate of functional decline in ALS. (**A**) Pearson correlation between protein slope and ALSFRS-R slope for each of the 7,550 aptamers in patients with serial plasma draws (n = 229), plotted against -log_10_ p. Slopes were estimated using linear mixed-effects models. The dashed line marks an FDR of 5%. Aptamers that fell (blue) or rose (red) with faster decline are colored and selected proteins are labeled. (**B**) Percentage of the differential plasma proteins that were also progression markers, shown for those that were higher (7 of 37) and lower (16 of 29) in patients with ALS than in controls. The OR and *p*-value are from Fisher’s exact test.

The 72 progression markers were therefore often proteins that were lower in patients than in controls, whereas several of the most elevated muscle proteins did not change with the rate of decline.

### Differential plasma proteins and progression markers replicate in an independent ALS cohort

To test whether these findings depended on the cohort or the SomaScan platform, we examined plasma from the Target ALS Natural History Study, an independent cohort profiled by tandem mass tag (TMT) mass spectrometry (57 patients with ALS and 43 controls) and by Olink proximity extension assay (60 patients and 56 controls; Table S5) (*24*). Of the 66 differential plasma proteins, 13 were measured by mass spectrometry and 42 by Olink. After adjustment for sex and age, 11 of 13 (85%) changed in the same direction on mass spectrometry (binomial *p* = 0.022) and 34 of 42 (81%) on Olink (*p* = 7 x 10^-5^). Their fold changes were strongly correlated with those in Answer ALS (r = 0.95 and 0.68, respectively), compared with r = 0.27 and r = 0.30 across all proteins shared by the two cohorts on each platform (Fig. 3, A and B; Table S6). On Olink, the muscle proteins ACTN2, CA3, MYL11, myosin-binding protein C1 (MYBPC1), myosin light chain 3 (MYL3), and cysteine and glycine-rich protein 3 (CSRP3) were elevated in ALS at an FDR of 5%, and MSTN was reduced (*p* = 0.007).

**Figure 3.**
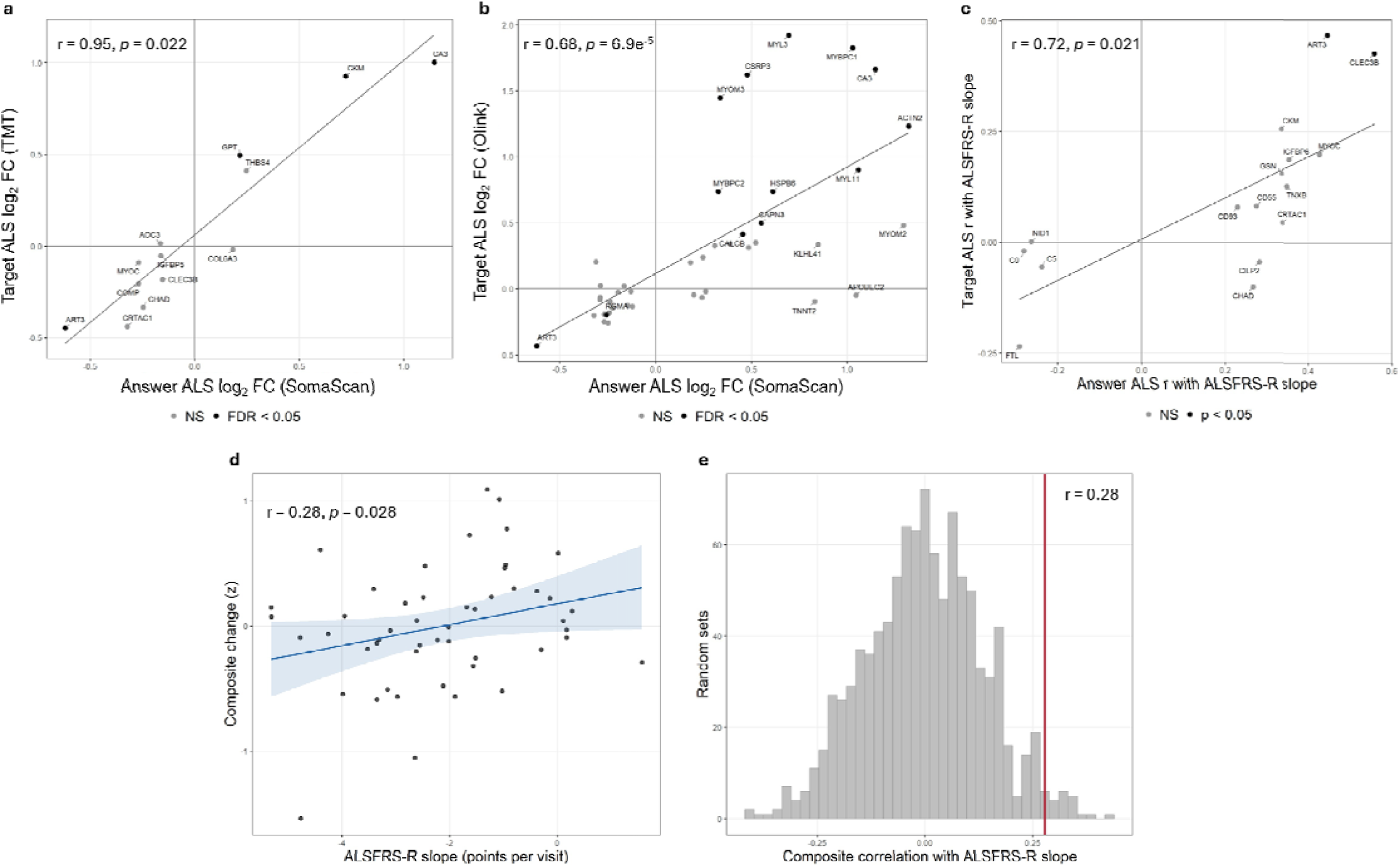
Differential plasma proteins and progression markers replicate in an independent ALS cohort. (**A** and **B**) Log_2_ fold change (FC) between patients with ALS and controls in Target ALS plotted against Answer ALS for the differential plasma proteins measured by tandem mass tag (TMT) mass spectrometry (**A**, n = 13) and by Olink (**B**, n = 42). Black points indicate an FDR below 5% in Target ALS and gray points were not significant (NS). (**C**) Correlation between protein slope and ALSFRS-R slope in Target ALS plotted against that in Answer ALS for the 16 progression markers measured by mass spectrometry. Black points indicate *p* < 0.05 in Target ALS. Lines in (**A**) to (**C**) show the linear fit. (**D**) Change in the composite of progression markers plotted against ALSFRS-R slope in Target ALS patients (n = 50), with each marker’s direction fixed from Answer ALS. The line and shaded band show the linear fit and its 95% confidence interval (CI). (**E**) Correlation with ALSFRS-R slope for 1,000 composites of randomly selected non-marker proteins measured on the same platform. The red line marks the composite of progression markers.

We then tested the progression markers in the 50 Target ALS patients with two or more mass spectrometry samples and repeated ALSFRS-R scores, using the same two-stage design with visit number as the time axis. Of the 60 progression markers with a unique gene symbol, 16 were measured in enough patients to estimate slopes. Thirteen (81%) of these were associated with the rate of decline in the same direction as in Answer ALS (binomial *p* = 0.021), and their correlations were closely related to those in Answer ALS (r = 0.72, *p* = 0.001), compared with ρ = 0.13 across all 563 shared proteins (Fig. 3C; Table S7). CLEC3B (r = 0.43, *p* = 0.002) and ART3 (r = 0.47, *p* = 0.002) replicated directly. Change in a composite of these progression markers, with each marker’s direction fixed from Answer ALS, also tracked decline (r = 0.28, 95% CI 0.03 to 0.50, permutation *p* = 0.028; Fig. 3D). It did so more closely than composites of randomly selected non-marker proteins measured on the same platform (empirical *p* = 0.022; Fig. 3E) and was robust to the removal of any single marker (r = 0.19 to 0.33; Fig. S4).

The muscle-dominated differential plasma proteins and the progression markers therefore replicated in an independent cohort on two platforms that do not depend on aptamer binding.

### A patient iPSC-derived motor neuron map separates plasma proteins of neuronal and non-neuronal origin

To test whether these plasma changes reflect motor neuron biology, we built a motor neuron expression map of the plasma proteome using patient-derived iMNs (Fig. 4A). Of the 7,550 aptamers, 7,175 mapped to an annotated gene, representing 6,301 unique plasma proteins (Table S8). For each, we asked whether it was detected in iMNs by RNA-sequencing and mass spectrometry, and whether its promoter was accessible on ATAC-sequencing. Plasma proteins detected at both transcript and protein level were classed as expressed and divided into promoter-supported when the promoter was also accessible (n = 1,459, 20.3%) and promoter-unconfirmed when it was not (n = 408, 5.7%). Those detected at the transcript level only were classed as transcript only (n = 3,937, 54.9%), those detected at the protein level only as protein only (n = 10, 0.1%), and those detected at neither level as not expressed (n = 1,361, 19.0%; Fig. 4B; Table S9). This class was stable across protein detection thresholds from 30% to 90% of iMN lines, varying by fewer than ten plasma proteins (Fig. S5). The large transcript-only class reflected the limited coverage of mass spectrometry, which did not measure 94.5% of these plasma proteins at all, whereas RNA-sequencing measured nearly every gene on the panel.

**Figure 4.**
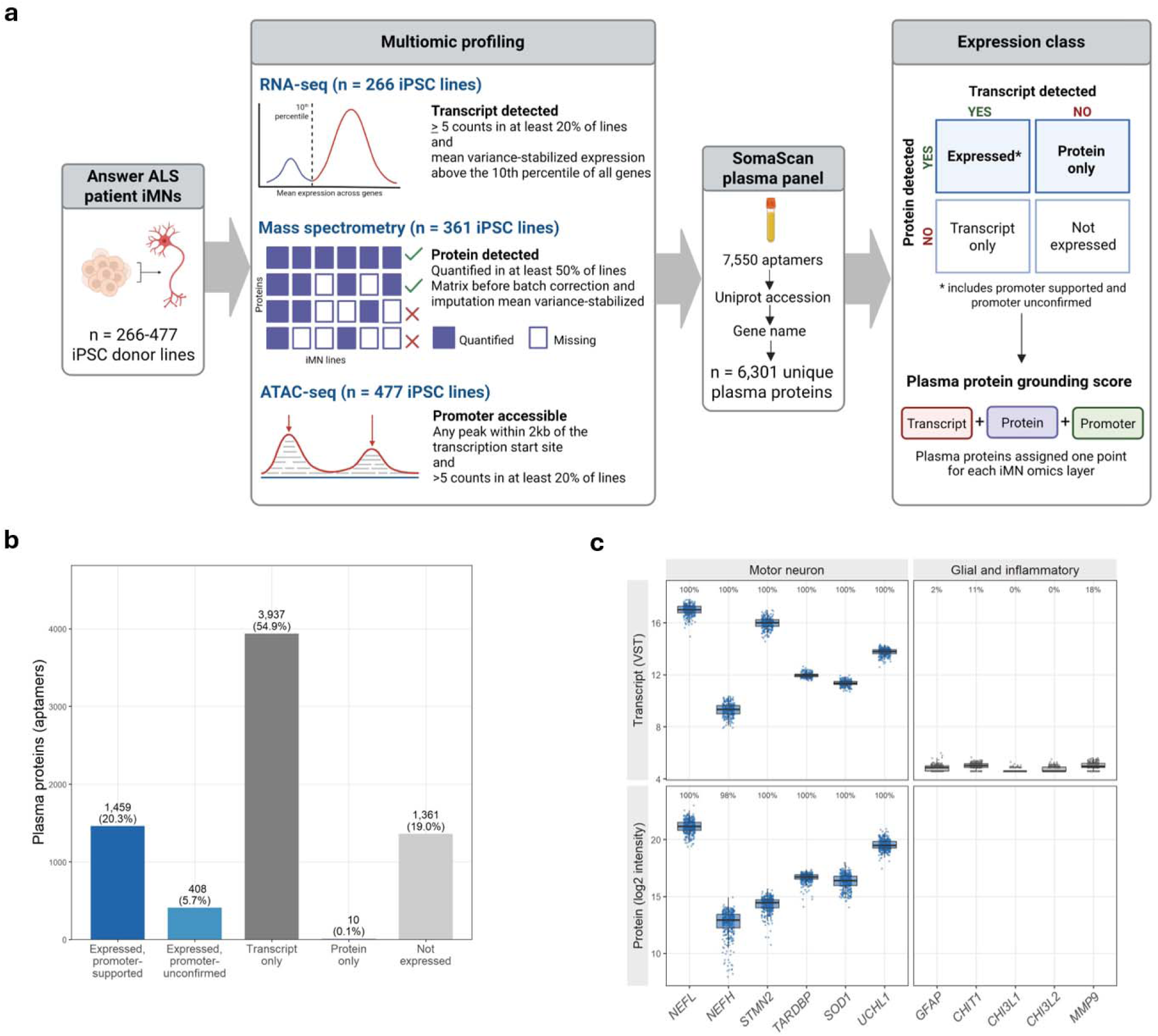
A patient iPSC-derived motor neuron map separates plasma proteins of neuronal and non-neuronal origin. (**A**) Construction of the motor neuron expression map from patient iMNs profiled by RNA-sequencing (n = 266 lines), mass spectrometry (n = 361 lines), and ATAC-sequencing (n = 477 lines). The 7,550 SomaScan aptamers were mapped by UniProt accession to 6,301 unique plasma proteins, and each was classed by whether its transcript and protein were detected in iMNs and, when expressed, by whether its promoter was accessible. The grounding score assigned each plasma protein one point for each of these three omics layers. (**B**) Number and percentage of plasma proteins in each class of the map, counted at the aptamer level (n = 7,175). (**C**) Transcript and protein abundance in iMNs for canonical ALS and neurodegeneration-associated proteins (blue) and for glial and inflammatory markers (gray). Points show individual iMN lines and boxes the median and interquartile range. The percentage of iMN lines in which each transcript or protein was detected is given above each plot. None of the glial and inflammatory markers was quantified by mass spectrometry. Abbreviations: iPSC: induced pluripotent stem cell, iMN: iPSC-derived motor neuron, VST: variance-stabilized transformation.

We next sought to validate the map using proteins with well-characterized expression in motor neurons and glia. The canonical ALS and neurodegeneration-associated proteins neurofilament light and heavy chain (NEFL, NEFH), stathmin-2 (STMN2), TAR DNA-binding protein (TARDBP), superoxide dismutase 1 (SOD1), and ubiquitin carboxy-terminal hydrolase L1 (UCHL1) were all detected in iMNs at both transcript and protein level (Fig. 4C). In contrast, neither transcript nor protein was detected for the glial and inflammatory markers glial fibrillary acidic protein (GFAP), chitotriosidase 1 (CHIT1), chitinase-3-like proteins 1 and 2 (CHI3L1, CHI3L2), and matrix metalloproteinase-9 (MMP9). Promoter chromatin was nonetheless accessible for all of these except CHI3L2, confirming that promoter accessibility alone does not indicate expression.

Overall, the map correctly separates known motor neuron proteins from glial and inflammatory proteins and can therefore be used to test whether a given plasma protein is expressed by motor neurons.

### Motor neuron proteins are underrepresented among plasma proteins that track decline or distinguish ALS from controls

We then asked whether plasma proteins expressed in iMNs were more likely than others to track functional decline. The mean absolute correlation between protein slope and ALSFRS-R slope was 0.071 for both classes of expressed plasma proteins, 0.066 for transcript-only, and 0.067 for not expressed plasma proteins (Fig. 5A). Each plasma protein was given a grounding score from 0 to 3 based on the number of omics layers with evidence in iMNs. Grounding score was associated with tracking (β = 0.0022 per point, *p* = 0.0007) but accounted for only 0.16% of the variance. Expressed plasma proteins also had a higher baseline signal than other plasma proteins (11.0 versus 10.2 log_2_ RFU). Plasma proteins with a higher signal are measured with less noise and therefore track decline more closely. After adjustment for baseline signal and its coefficient of variation, grounding score was no longer associated with tracking (*p* = 0.15, permutation *p* = 0.13), and the difference between expressed and transcript-only plasma proteins disappeared (*p* = 3.8 x 10^-9^ before and *p* = 0.42 after adjustment; Fig. 5B). The same was true using external blood concentration estimates from the Human Protein Atlas (*25*) in place of SomaScan signal, with no association between grounding score and tracking before (*p* = 0.29) or after (*p* = 0.57) adjustment among 3,181 plasma proteins (Fig. S6).

**Figure 5.**
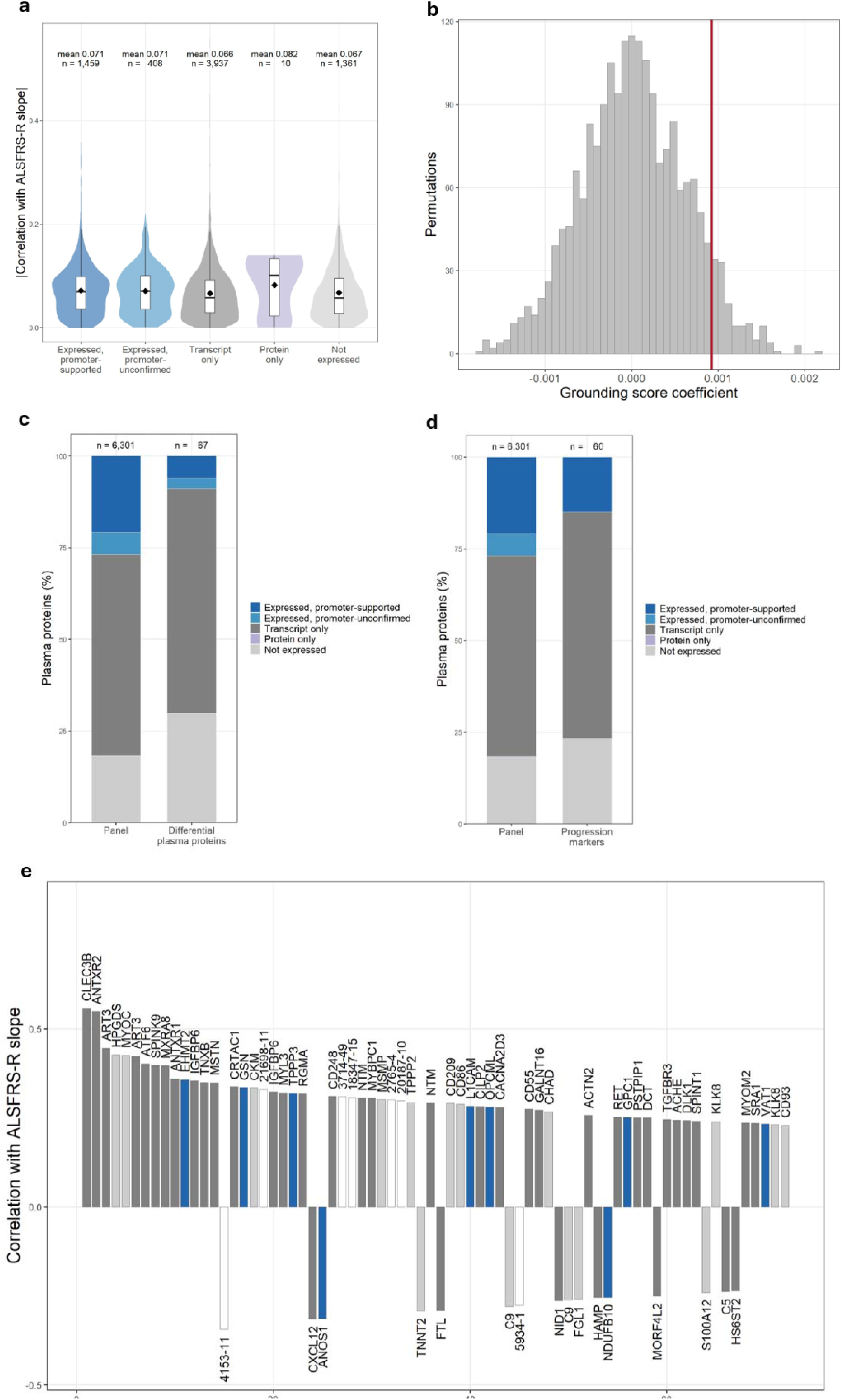
Motor neuron proteins are underrepresented among plasma proteins that track decline or distinguish ALS from controls. (**A**) Absolute correlation between protein slope and ALSFRS-R slope for plasma proteins in each class of the motor neuron expression map. Violins show the distribution, boxes the median and interquartile range, and diamonds the mean. The mean and number of plasma proteins are given above each class. (**B**) Grounding score coefficients from 2,000 permutations of grounding score across aptamers, after adjustment for baseline signal and its coefficient of variation. The red line marks the observed coefficient. (**C** and **D**) Percentage of plasma proteins in each class for the differential plasma proteins (**C**, n = 67) and the progression markers (**D**, n = 60), each compared with the panel (n = 6,301). (**E**) Correlation with ALSFRS-R slope for the 72 progression markers, ranked by strength of association and colored by class. Aptamers without an annotated gene are shown in white and labeled with their SomaScan sequence identifier.

Of the 67 differential plasma proteins that could be classed, only six (9%) were expressed in iMNs, compared with 27% of the panel (OR = 0.26, 95% CI 0.09 to 0.61, *p* < 0.001), and 20 (30%) were not expressed at all, including the muscle proteins apolipoprotein B mRNA editing enzyme catalytic subunit 2 (APOBEC2), CKM, TNNI2, cardiac troponin T (TNNT2), and CSRP3 (Fig. 5C). Of the 60 progression markers that could be classed, only nine (15%) were expressed in iMNs (OR = 0.48, 95% CI 0.21 to 0.98, *p* = 0.04), while 37 (62%) were transcript only and 14 (23%) were not expressed (Fig. 5D). The distribution of the differential plasma proteins across all five classes differed from the panel (Fisher *p* = 0.004), whereas that of the progression markers did not (*p* = 0.12). The nine progression markers most strongly associated with decline, including CLEC3B, ANTXR2, and ART3, were all transcript only or not expressed in iMNs, and the first expressed progression marker, euchromatic histone-lysine N-methyltransferase 2 (EHMT2, r = 0.36), ranked tenth (Fig. 5E; Table S10).

Plasma proteins expressed in iMNs are therefore underrepresented among both sets and expression in iMNs does not predict whether a plasma protein tracks functional decline in ALS.

### ALS progression markers enter plasma by secretion or shedding and are not specific to any tissue

We next asked how the progression markers reach plasma and in which tissues they are found. Based on UniProt subcellular localization (*26*), 27 (45%) of the 60 progression markers were secreted and 18 (30%) were membrane proteins that reach plasma by shedding from the cell surface. Together these accounted for 75% of progression markers compared with 48% of the panel. Only 12 (20%) progression markers were intracellular proteins that reach plasma when cells are damaged, compared with 43% of the panel (OR = 0.30, 95% CI 0.14 to 0.57, *p* = 8 x 10^-5^; Fig. 6A; Table S11), and none of these was a neuronal protein. They comprised three muscle proteins (CKM, ACTN2, MYOM2), seven broadly expressed proteins, including EHMT2 and ferritin light chain (FTL), and two tissue-specific proteins, including HPGDS (Fig. S7A). Three further progression markers, the sarcomeric proteins MYL3, MYBPC1, and TNNT2, had no UniProt subcellular localization and were not assigned a route. In contrast, many of the differential plasma proteins were sarcomeric and cytosolic muscle proteins, which are established serum markers of muscle fiber damage (*27–29*), although intracellular proteins as a whole were not significantly enriched among them (30% versus 43% of the panel, OR = 0.60, *p* = 0.08). Of the nine progression markers expressed in iMNs, only two, L1 cell adhesion molecule (L1CAM) and opioid-binding protein/cell adhesion molecule (OPCML; both r = 0.28), were consistent with shedding from the neuronal surface (Fig. 6B).

**Figure 6.**
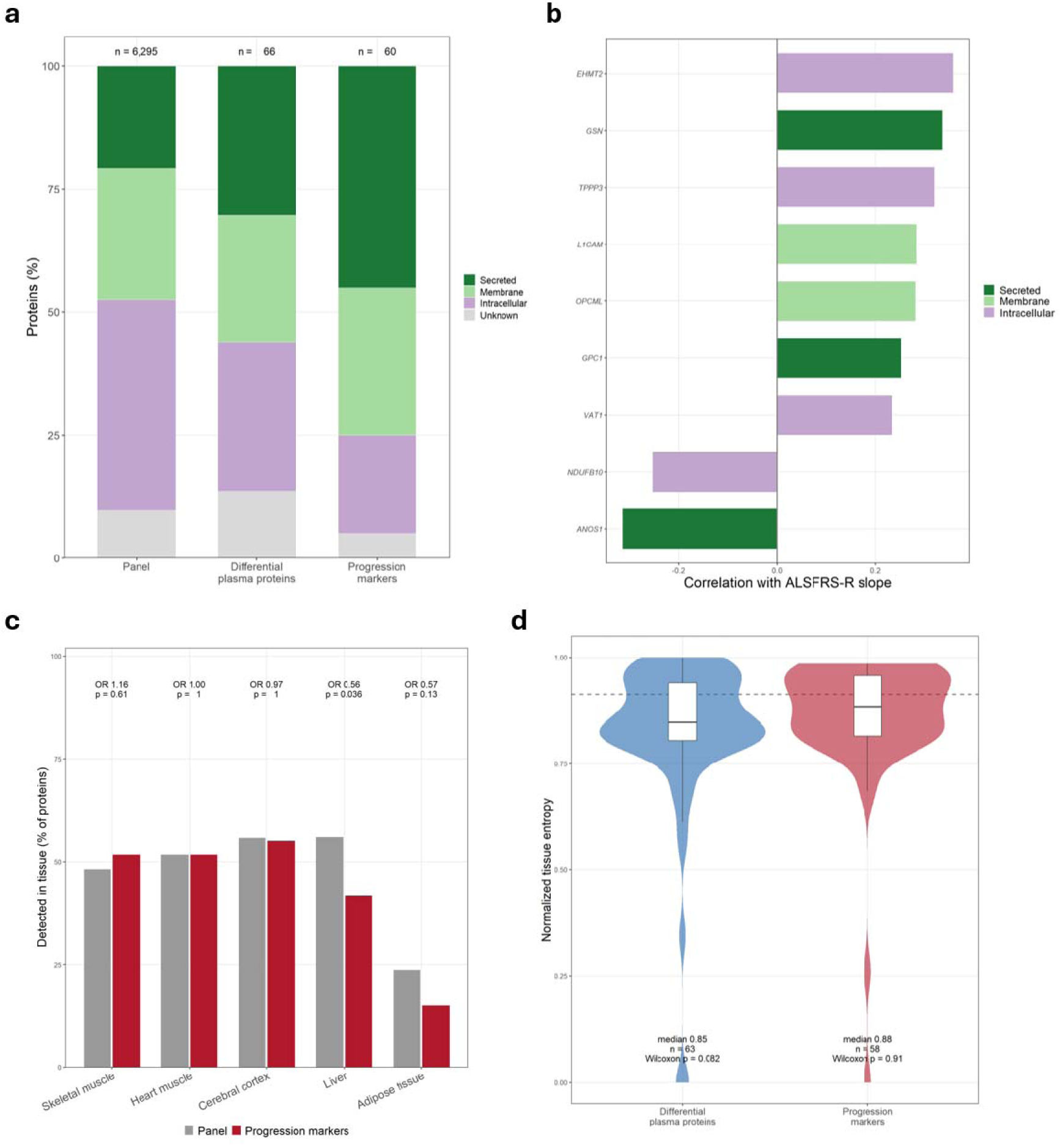
ALS progression markers enter plasma by secretion or shedding and are not specific to any tissue. (**A**) Percentage of secreted, membrane, and intracellular proteins among the panel (n = 6,295), the differential plasma proteins (n = 66), and the progression markers (n = 60), based on UniProt subcellular localization. (**B**) Correlation with ALSFRS-R slope for the nine progression markers expressed in iMNs, colored by route to plasma. (**C**) Percentage of the progression markers (red) and of the panel (gray) detected in each tissue. ORs and *p*-values compare the progression markers with the panel. (**D**) Normalized tissue entropy across 59 human tissues for the differential plasma proteins (n = 63) and the progression markers (n = 58). Violins show the distribution and boxes the median and interquartile range. The dashed line marks the median of the panel, and *p*-values are from Wilcoxon tests against the panel.

We then assessed the distribution of each progression marker across 59 human tissues using concordR (*30*), which combines protein evidence from the Human Protein Atlas (*31*), GTEx (*32*), and PaxDb (*33*). Unlike the differential plasma proteins, progression markers were not enriched for detection in skeletal muscle (52% versus 48% of the panel, OR = 1.16, *p* = 0.61) and were detected in cerebral cortex and heart at the same rate as the panel, with only a depletion in liver (42% versus 56%, OR = 0.56, *p* = 0.04; Fig. 6C). Progression markers were also as evenly spread across tissues as the panel, with a median normalized entropy of 0.88 compared with 0.91 (Wilcoxon *p* = 0.91), where a value of 1 indicates equal presence in all tissues (Fig. 6D; Table S12). Only 2 of the 58 progression markers with tissue evidence were concentrated in a single tissue. Cerebral cortex ranked first for 23 (40%) progression markers, but it ranked first for a similar 37% of the panel (OR = 1.12; *p* = 0.68; Fig. S7B), reflecting the breadth of protein detection in cortex across these atlases.

The differential plasma proteins are therefore dominated by muscle proteins that leak from damaged fibers, whereas the progression markers are secreted or shed proteins spread across many tissues.

### Serum NfL is prognostic at baseline but does not track decline over time

To test whether our approach could detect neuronal release in plasma, we next examined NfL, the established blood marker of neuroaxonal injury in ALS (*7, 10*). On SomaScan, the NEFL aptamer did not differ between patients and controls (*p* = 0.50) and was not associated with the rate of decline, either as a change over time (n = 229, r = -0.03, *p* = 0.62) or as a baseline level (n = 242, r = 0.06, *p* = 0.32; Fig. 7A). In contrast, change in CLEC3B and ANTXR2 tracked decline, and baseline CLEC3B predicted it after adjustment for baseline ALSFRS-R (r = 0.15, *p* = 0.027; Fig. 7A), showing that both designs could detect a true signal. To determine whether this reflected NfL biology or the NEFL aptamer, we used serum NfL measured by immunoassay in 740 patients from the same cohort. In the 241 patients with both measurements at the same visit, the aptamer agreed poorly with the immunoassay (r = 0.08), and within-patient change on the two platforms was uncorrelated (r = -0.02; Fig. 7B; Table S13).

**Figure 7.**
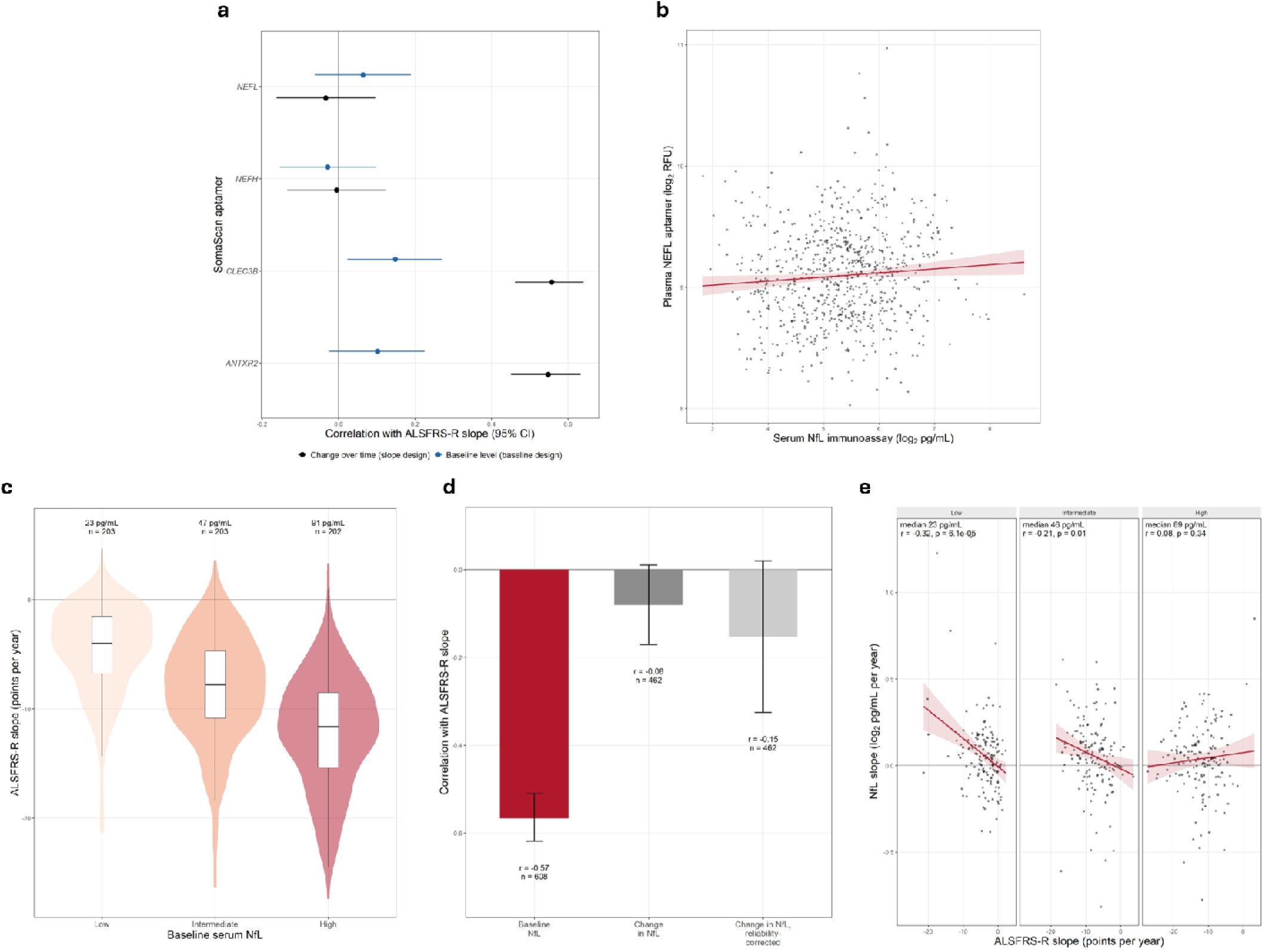
Serum NfL is prognostic at baseline but does not track decline over time. (**A**) Correlation with ALSFRS-R slope for the NEFL, NEFH, CLEC3B, and ANTXR2 aptamers on SomaScan, shown for change over time (black, n = 229) and for baseline level (blue, n = 242). Points and bars show the correlation and its 95% CI. (**B**) Plasma NEFL aptamer signal plotted against serum NfL measured by immunoassay in the 763 draws from 241 patients in which both were measured at the same visit. (**C**) ALSFRS-R slope in patients with low (n = 203), intermediate (n = 203), and high (n = 202) baseline serum NfL. The median baseline NfL is given above each group. Violins show the distribution and boxes the median and interquartile range. (**D**) Correlation with ALSFRS-R slope for baseline NfL (n = 608), change in NfL (n = 462), and change in NfL after correction for the reliability of the slope estimates. Error bars show the 95% CI. (**E**) NfL slope plotted against ALSFRS-R slope in patients with low, intermediate, and high baseline NfL. Lines and shaded bands in (**B**) and (**E**) show the linear fit and its 95% CI. Abbreviations: RFU: relative fluorescent units.

Measured by immunoassay, baseline serum NfL strongly predicted the subsequent rate of decline (n = 608, r = -0.57, *p* = 6 x 10^-53^), independently of baseline ALSFRS-R score. Patients in the lowest, middle, and highest thirds of baseline NfL (median 23, 47, and 91 pg/mL) lost 4.7, 8.1, and 12.0 ALSFRS-R points per year, respectively (Kruskal-Wallis *p* = 2 x 10^-41^; Fig. 7C). In contrast, within-patient change in NfL was only weakly related to the rate of decline in the 462 patients with two or more NfL measurements (r = -0.08, *p* = 0.08), and remained weak after correction for the reliability of the slope estimates (r = -0.15; Fig. 7D). When patients were grouped by baseline NfL, change in NfL tracked decline in the lowest third (r = -0.32, *p* = 6 x 10^-5^) and middle third (r = -0.21, p = 0.01), but not in the highest (r = 0.08, *p* = 0.34; Fig. 7E). Grouping on a single baseline measurement can produce this pattern through regression to the mean, so we re-estimated NfL slopes from follow-up samples alone in the 275 patients with three or more samples. Change in NfL still tracked decline in the lowest third (r = -0.23, *p* = 0.02) but not in the middle or highest thirds (highest r = 0.08, *p* = 0.51), and the difference between the lowest and highest thirds persisted (*p* = 0.04; Fig. S8).

A single baseline NfL measurement therefore predicts how quickly a patient will decline, whereas change in NfL tracked decline only in patients whose NfL was low at baseline. The absence of NEFL from the progression markers likely reflects how NfL is measured by the SomaScan aptamer in plasma and not NfL biology.

### Change in the progression markers tracks decline within strata of baseline NfL

We next asked whether the progression markers carried information about decline that NfL did not. Change across the 72 progression markers was summarized for each patient as a composite score. The markers were selected in the same patients, so the composite was cross-fitted, with markers selected in one half of the cohort and scored in the other. In the 229 patients with serial SomaScan and NfL measurements, change in the cross-fitted composite was strongly associated with the rate of decline (r = 0.475, *p* = 3 x 10^-14^), whereas change in NfL was not (r = 0.02, *p* = 0.74). The difference between the two correlations was 0.48 (95% CI 0.28 to 0.67). Baseline NfL and change in NfL together explained 36% of the variance in the rate of decline, and adding the composite increased this to 45% (*p* = 1 x 10^-9^).

Baseline NfL is measured at enrolment and is already used to stratify patients in clinical trials (*13*), so we repeated this comparison within thirds of baseline NfL (median 21, 41, and 71 pg/mL). Change in the composite tracked decline in every third (r = 0.49, 0.37, and 0.39 from the lowest to the highest). Change in NfL was weakly associated with decline in the lowest and middle thirds (r = 0.33 and 0.23) and was not associated with it in the highest (r = 0.07; Fig. 8A). The difference between the two was largest in the highest third (Δr = 0.32, *p* = 0.061), where patients declined fastest, and was in the same direction in the lowest (Δr = 0.17, *p* = 0.34) and middle thirds (Δr = 0.15, p = 0.29).

**Figure 8.**
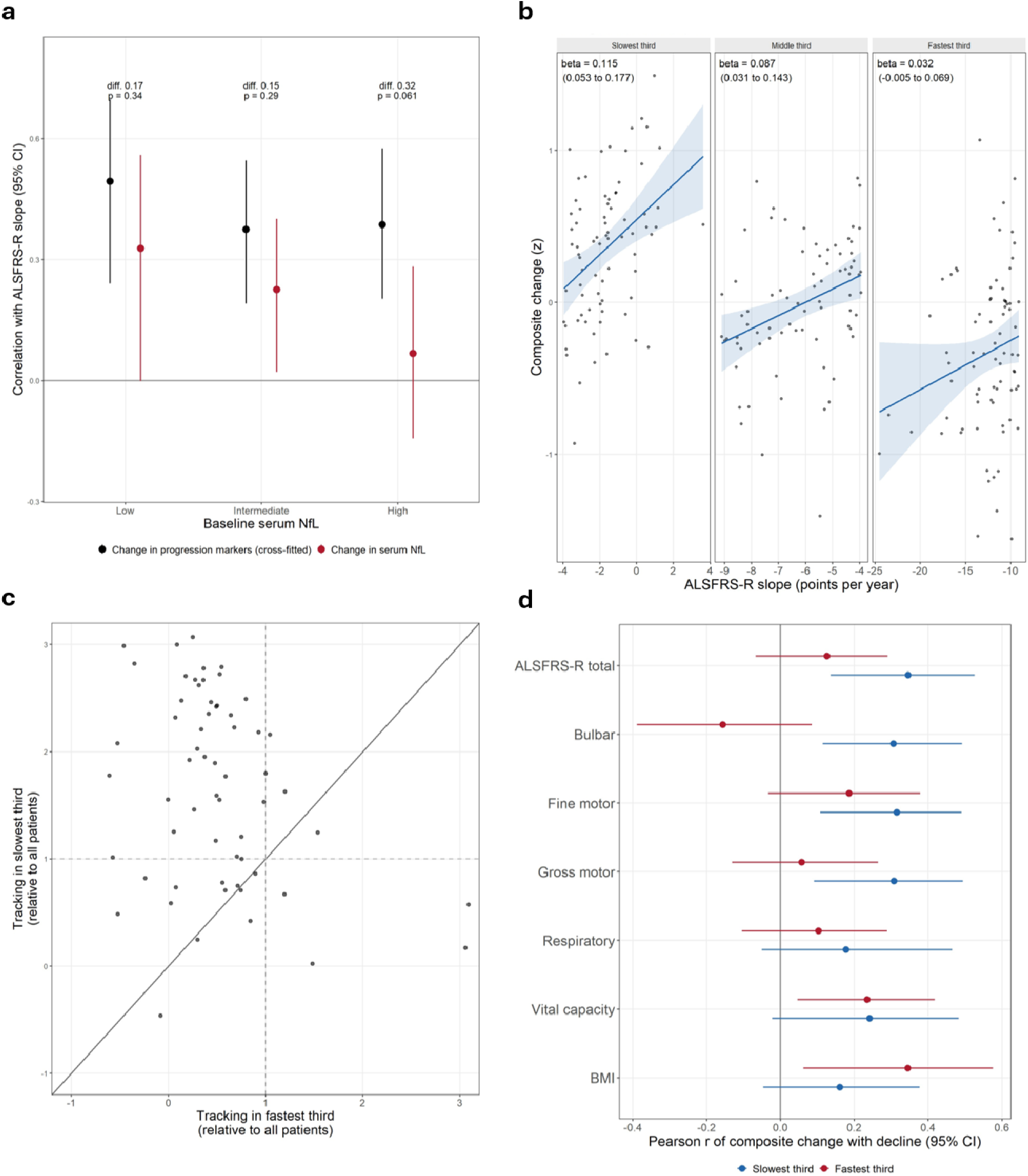
Change in the progression markers tracks decline across baseline NfL but not in the fastest progressing patients. (**A**) Correlation with ALSFRS-R slope for change in the cross-fitted composite of progression markers (black) and for change in serum NfL (red) in patients with low, intermediate, and high baseline NfL (n = 229). Points and bars show the correlation and its 95% CI. The difference between the two correlations and its *p*-value are given above each group. (**B**) Change in the composite plotted against ALSFRS-R slope in the slowest, middle, and fastest thirds of patients by rate of decline. Lines and shaded bands show the linear fit and its 95% CI, and the coefficient (beta) is given with its 95% CI. (**C**) Tracking of each of the 72 progression markers in the slowest third plotted against that in the fastest third, each relative to tracking in all patients. Dashed lines mark tracking equal to that in all patients, and markers above the diagonal tracked decline less closely in the fastest than in the slowest third. (**D**) Pearson correlation of change in the composite with decline in ALSFRS-R total score, each ALSFRS-R subdomain, vital capacity, and body mass index (BMI) in the slowest (blue) and fastest (red) thirds. Points and bars show the correlation and its 95% CI.

Change in the progression markers therefore tracked functional decline whether baseline NfL was low, intermediate, or high, whereas change in NfL did not.

### The progression markers do not track decline in the fastest progressing patients

To test whether the progression markers tracked decline equally across patients, we divided the 229 patients into thirds by their rate of decline (median -11.7, -5.9, and -1.9 ALSFRS-R points per year). Change in the composite was associated with the rate of decline in the slowest third (0.115 composite units per ALSFRS-R point per year, 95% CI 0.053 to 0.177) but not in the fastest third (0.032, 95% CI -0.005 to 0.069, interaction *p* = 0.026; Fig. 8B). The cross-fitted composite showed the same difference (0.016 versus 0.078, *p* = 0.038), and 60 of the 72 progression markers tracked decline less closely in the fastest than in the slowest third (Fig. 8C). The difference was driven by the fastest progressors. Whether patients were divided into halves or quintiles, the coefficient in the slowest group remained between 0.06 and 0.08, whereas that in the fastest group fell from 0.022 in halves (*p* = 0.007 for the difference) to 0.004 in quintiles (Fig. S9A; Table S14). The fastest third had shorter follow-up than the slowest (1.2 versus 1.9 years) and included 43 of the 75 patients who died during follow-up. However, the difference persisted in patients followed for at least one year (*p* = 0.048), and adjusting for follow-up length reduced it by about a fifth. It also persisted after excluding protein and ALSFRS-R samples taken within six months of death (*p* = 0.013) and was in the same direction after excluding those taken within 12 months (*p* = 0.07; Fig. S9B).

In the fastest third, change in the composite did not track decline in any ALSFRS-R subdomain (r = -0.16 to 0.19), whereas in the slowest third it tracked decline in the bulbar, fine motor, and gross motor subdomains (r = 0.31 to 0.32) but not in the respiratory subdomain (r = 0.18; Fig. 8D; Table S14). It retained a weaker association with the decline in vital capacity (r = 0.24, 95% CI 0.05 to 0.42) and with the loss of body mass index (BMI; r = 0.35, 95% CI 0.06 to 0.58), similar to that in the slowest third (r = 0.24 and 0.16). Within the fastest third, tracking did not differ significantly between the 15 patients with bulbar onset and the 62 with limb or other onset (interaction p = 0.30; Table S14).

At enrolment, patients in the fastest third had a shorter symptom duration than those in the slowest (median 1.4 versus 3.3 years, FDR = 1 x 10^-6^) and a faster rate of decline before enrolment (ΔFS 0.59 versus 0.24 points per month, FDR = 7 x 10^-9^). They also had a higher baseline NfL (58.5 versus 24.4 pg/mL, FDR = 7 x 10^-15^), but similar baseline ALSFRS-R scores (median 37 versus 39.5) and proportions with bulbar onset (19% versus 16%; Table S15). Nine of the 17 C9orf72 expansion carriers were in the fastest third, compared with one in the slowest although the difference was not significant (12% versus 1%, FDR = 0.10). During follow-up, the fastest third also declined faster in every ALSFRS-R subdomain and vital capacity, and more often required gastrostomy (23% versus 8%) and non-invasive ventilation (64% versus 34%; Table S15). Baseline NfL identified patients in the fastest third more accurately than ΔFS (AUC = 0.79 versus 0.72) but change in the composite still tracked decline in the highest third of baseline NfL (r = 0.44) and of ΔFS (r = 0.37).

The progression markers therefore lost track of decline in the fastest progressing patients, as did change in NfL in patients with the highest baseline NfL.

## Discussion

The Answer ALS cohort is the largest collection of longitudinal clinical data, serial biospecimens, and patient iPSC lines from individuals with ALS assembled to date (*22, 23*). This enabled us to ask whether the plasma proteins that change as patients with ALS decline originate from motor neurons, and to compare how their change and change in NfL track decline. We show that plasma proteins that distinguish patients from controls are dominated by muscle proteins leaking from damaged fibers, whereas those whose change tracks the rate of decline are secreted or shed proteins found across many tissues. Both sets replicated in an independent cohort, and a motor neuron expression map built from the same patients showed that motor neuron expression does not predict whether a plasma protein tracks decline. Baseline serum NfL predicts how quickly patients decline but its change does not track decline, whereas change in the progression markers does so within every stratum of baseline NfL. Neither tracks decline in the fastest progressing patients.

The muscle-dominated differences between patients and controls are in line with recent large plasma proteomic studies. On SomaScan, Dergai et al. reported the largest increases in PDLIM3, TNNT2, and MYL11 and the largest decreases in ANTXR2 and ART3, with CLEC3B among their progression markers and a fall in MSTN (*16*). Muscle proteins were also prominent on Olink in patients and pre-symptomatic samples (*15, 34*). Our study extends these findings by showing that the proteins whose within-patient change tracks decline are largely distinct from those that best distinguish patients from controls, and by testing directly whether either set comes from motor neurons.

To date, the tissue origin of ALS plasma proteins has been inferred from tissue expression databases (*15, 16*). Dergai et al., for example, reported that CLEC3B and ANTXR2 are most highly expressed in microglia and Schwann cells, respectively (*16*). In contrast, we tested motor neuron origin directly for each plasma protein using iMNs from patients in the same cohort, although our map is limited to motor neurons and cannot exclude an origin in these or other cell types. This raises two considerations. First, a plasma protein that tracks decline in a neurodegenerative disease does not need to come from the degenerating neurons. The strongest progression markers, CLEC3B, ANTXR2, and ART3, were not expressed or were transcript only in iMNs, and several, including CLEC3B and CRTAC1, have been linked to aging, longevity, and joint degeneration in population studies (*35, 36*). Their decline in plasma may therefore reflect the systemic consequences of ALS, such as denervation, immobility, weight loss, and hypermetabolism (*19*) on tissues throughout the body. ART3, which is highly expressed in skeletal muscle (*16*), was lower in patients and fell further with decline, suggesting that muscle contributes to the progression signal through proteins that fall as well as through proteins that leak. The insulin-like growth factor binding protein (IGFBP6) was also a progression marker, and the related IGFBP2 has been associated with survival in ALS (*37*). This concerns where the plasma signal of progression comes from and not where the disease begins. Whether degeneration starts in the motor cortex (*38*) or the distal axon and neuromuscular junction (*39*) plasma proteins that track decline could reflect the consequences of motor neuron loss for muscle and other tissues. Second, the differential plasma proteins and the progression markers reflect different processes. Muscle proteins such as CA3, TNNI2, and MYL11 were the most elevated in patients but did not change with the rate of decline, consistent with ongoing leak from denervated muscle that is present throughout the disease (*29, 40*). In contrast, the progression markers mostly fell as patients declined. Plasma levels also depend on how a protein is released and not only on its expression, as shown for FGF23, which is increased in ALS muscle but not in plasma (*41*). Plasma proteins chosen for their difference from controls are therefore not necessarily suited to monitoring progression.

Unexpectedly, the SomaScan NEFL aptamer did not differ between patients and controls and agreed poorly with NfL measured by immunoassay at the same visits. NEFL was measured in plasma and NfL in serum, but NfL levels in the two differ only slightly (*42, 43*), which cannot account for the low correlation. The SomaScan NEFL aptamer has also shown no agreement with immunoassay NfL in plasma (*44*), whereas it agrees closely with immunoassay in CSF (*45*) where NfL is far more abundant. These results suggest that the aptamer does not reliably measure NfL at the concentrations found in blood, and NEFL results from SomaScan plasma data should be treated with caution. This is a limitation of a single aptamer and not of the platform broadly, as the progression markers identified on SomaScan replicated in mass spectrometry in Target ALS. Measured by immunoassay, baseline NfL was strongly prognostic, consistent with previous studies (*9, 10, 12*). In contrast, change in NfL over time was only weakly related to the rate of decline, even after correction for the reliability of the slope estimates. This is consistent with the relative stability of NfL within patients (*9, 10*) and suggests that NfL largely reflects each patient’s overall rate of neuroaxonal loss and not the decline in function as it accumulates. The exception was patients with low baseline NfL, in whom change in NfL did track decline.

Our findings have direct implications for clinical trials and patient care. NfL is already used to stratify patients and as a pharmacodynamic endpoint (*13, 14*), but our results suggest that change in NfL is a poor measure of whether functional decline is slowing in most patients. Change in the progression markers tracked decline irrespective of baseline NfL and explained decline beyond it. This supports a complementary approach in which baseline NfL is used to stratify patients at enrolment and a panel of systemic plasma proteins is used to monitor decline during follow-up. The replication of progression markers on mass spectrometry, including CLEC3B and ART3 individually, suggests that a panel would not be restricted to a single platform. Plasma proteins and ALSFRS-R were measured over the same period, so our analyses show that the progression markers track decline as it occurs and not that they predict future decline. Whether they respond to treatment, however, remains to be tested in trial samples.

The progression markers did not track decline in the fastest progressing patients. This was not explained by shorter follow-up or by samples collected close to death and extended to every ALSFRS-R subdomain. Change in NfL similarly failed to track decline in patients with the highest baseline NfL, who overlapped with the fast progressors. Those patients could be recognized at enrolment by a short symptom duration and high baseline NfL and included about half of the C9orf72 expansion carriers. We interpret this not as a limitation of the markers but as evidence that the process driving decline in the fastest progressors is not reflected in change in either systemic or neuronal blood markers. The retained association with the decline in vital capacity and the loss of BMI suggests that respiratory and metabolic change in these patients may still be captured in plasma, whereas the rapid loss of limb and bulbar function is not. Proteomic differences between fast and slow progressors in longitudinal CSF (*21*) and changes in circulating immune cells (*18*) have been linked to rapid progression, so markers of decline in these patients may need to be tested in CSF or cells. Trials that monitor all patients with the same blood biomarkers may therefore be unable to detect a treatment effect in fast progressors. Cohorts with denser sampling earlier in the disease will be needed to identify markers of their decline.

Several further limitations should be considered. First, iMNs are immature relative to adult spinal motor neurons and their expression is influenced by differentiation and line effects (*46, 47*). Expression in iMNs may therefore differ from that in adult motor neurons, although iMNs expressed all of the canonical motor neuron proteins we tested. Second, the progression markers were identified on SomaScan and only 16 were measured in mass spectrometry in Target ALS, which also lacked visit dates and was too small to compare fast and slow progressors. The inability to track fast progressors therefore rests on a single cohort and requires replication. Third, patients who progress slowly are more likely to enroll later in their disease, which may contribute to the longer symptom duration of the slowest third.

Overall, we identify plasma proteins whose within-patient change tracks the rate of functional decline in ALS, replicate them in an independent cohort, and show using patient iMNs that they originate outside motor neurons. Change in these systemic proteins tracked decline across the range of baseline NfL, whereas change in NfL did not. This positions baseline NfL for prognosis and stratification and systemic plasma proteins for monitoring, an approach that can be applied directly to the design of clinical trials. We also identify the fastest progressing patients as a group whose decline is tracked by neither, highlighting the need for precision biomarker studies designed around these patients.

## Materials and Methods

### Study design

The objective of this study was to identify plasma proteins whose within-patient change tracks the rate of functional decline in ALS, to test whether they originate from motor neurons, and to compare them with serum NfL. We used all participants available in the Answer ALS and Target ALS public data releases who met the data requirements of each analysis. As the data were generated previously (*22–24*) there was no further randomization or blinding.

### Cohorts

Answer ALS recruited individuals with sporadic and familial ALS and age-matched neurologically unimpaired controls across eight US neuromuscular centers (*22*). Sex was self-reported (Table S1). Functional impairment was assessed at each visit using the ALSFRS-R total score (*2*). Time was defined as years since screening. Disease duration was calculated as the difference between age at first peripheral blood mononuclear cell (PBMC) collection and age at symptom onset. The Target ALS Global Natural History Study is a multi-center longitudinal cohort in which plasma was profiled by TMT mass spectrometry and Olink (Table S5) (*24*). Visit dates were not available in the clinical release, so time was represented by visit number. All participants provided written informed consent, and study procedures were approved by the institutional review boards of each participating site and adhered to the principles of the Declaration of Helsinki (*22, 24*). No separate ethics approval was required as this study used publicly available data.

### Plasma proteomics and serum NfL

Plasma was profiled using the SomaScan v4.1 assay, which measures approximately 7,500 aptamers using slow off-rate modified aptamers (*48*). Data were normalized by SomaLogic using adaptive normalization by maximum likelihood, and relative fluorescent units (RFU) were log_2_-transformed. The 7,550 aptamers with an annotated UniProt accession were analyzed separately and identified by their SomaScan sequence identifier. The baseline sample was each participant’s earliest sample. Serum NfL was measured by Answer ALS on the Siemens Atellica immunoassay platform, reported in pg/mL, and log_2_-transformed, giving 1,655 draws from 740 patients with ALS.

### Motor neuron expression map

iPSCs were generated from patient PBMCs and differentiated into iMNs, which were profiled on day 32 by RNA-sequencing, label-free mass spectrometry, and ATAC-sequencing (*22, 23*). Each aptamer was mapped from its UniProt accession to a gene. A plasma protein was considered detected at the transcript level if its gene had five or more counts in at least 20% of iMN lines and a mean variance-stabilized expression above the 10^th^ percentile across all genes. It was detected at the protein level if it was quantified in at least 50% of iMN lines in the matrix before batch correction and imputation. Its promoter was considered accessible if any ATAC-sequencing peak within 2 kb of its transcription start site had more than five counts in at least 20% of lines. Plasma proteins were assigned to the five classes of the map from these calls, with expressed plasma proteins classed as promoter-unconfirmed when no accessible promoter peak was identified. A grounding score from 0 to 3 was assigned to each plasma protein as the number of layers (transcript, protein, and accessible promoter) with supporting evidence in iMNs. Processing and validation are described in the Supplementary Methods.

### Differential plasma proteins

Differences in baseline plasma protein levels between patients with ALS and controls were assessed using linear models with empirical Bayes moderation in ‘limma’ (*49*), adjusting for sex and age. Plasma proteins with a Benjamini-Hochberg (*50*) FDR below 0.05 were considered differential plasma proteins. Within patients, each differential plasma protein was related to the baseline ALSFRS-R score and to disease duration by Pearson correlation, with FDR controlled across the 77 proteins for each.

### Longitudinal analyses

Within-patient change in plasma proteins was related to the rate of functional decline using a two-stage design (*51, 52*). In the first stage, each patient’s rate of decline was estimated from all dated ALSFRS-R scores in patients with two or more assessments (n = 663) using a linear mixed-effects model in ‘lme4’ with a fixed effect of time and participant-specific random intercepts and slopes, fit by restricted maximum likelihood (*53*). Each patient’s rate of decline was taken as the best linear unbiased predictor (BLUP) of their slope (*54*). Each patient’s rate of change in each plasma protein was estimated in the same way in patients with two or more dated draws and an ALSFRS-R slope (n = 229), and an aptamer was tested only if at least 20 patients contributed two or more measurements. In the second stage, each patient’s protein slope was related to their ALSFRS-R slope by Pearson correlation, with FDR controlled across the 7,550 aptamers. Aptamers with an FDR below 0.05 were considered progression markers. ALSFRS-R slopes are negative in decline, so a positive correlation indicates that faster decline was accompanied by a faster fall in plasma level. The two-stage design was compared with alternative methods by simulation. Cohorts of 229 patients were simulated from the visit schedules, variance components, and dropout estimated in Answer ALS, each with 1,000 plasma proteins of which five tracked decline, and the methods were compared on power and FDR. The methods were then compared in the cohort on the number of aptamers identified and their replication between random halves (Fig. S1 to S3; Supplementary Methods).

### Motor neuron expression and progression tracking

Progression tracking was defined as the absolute correlation between protein slope and ALSFRS-R slope. For the 7,175 aptamers that mapped to an annotated gene, tracking was regressed on grounding score, first without adjustment and then with adjustment for each aptamer’s mean baseline signal and its coefficient of variation. The adjusted coefficient was compared against a null distribution from 2,000 permutations of grounding score across aptamers. Enrichment of map classes among the differential plasma proteins and progression markers was assessed against the panel using Fisher’s exact test. Route to plasma and tissue distribution were assessed using concordR (v0.2.0) (*30*) as described in the Supplementary Methods.

### NEFL and NfL

The NEFL aptamer (SomaScan sequence identifier 10082-251) was examined in the slope design (n = 229) and in a baseline design in which ALSFRS-R slope was regressed on baseline NEFL (n = 242), with CLEC3B and ANTXR2 as positive controls. Agreement between NEFL and NfL was assessed by Pearson correlation in draws in which both were measured at the same visit. NfL was then examined in the same two designs. The observed correlation between NfL slope and ALSFRS-R slope was corrected for the reliability of the two slope estimates (*55*). Regression to the mean was tested using NfL slopes re-estimated from follow-up draws alone, as described in the Supplementary Methods.

### Composite score and comparison with NfL

The composite score was calculated by standardizing each marker slope across patients, multiplying it by the sign of the marker’s correlation with ALSFRS-R slope, and averaging across the 72 progression markers. The composite was also cross-fitted by were randomly dividing patients into two halves, selecting the 72 aptamers most strongly correlated with ALSFRS-R slope in one half, and scoring the composite in the other using the signs from the first. This was done in both directions and repeated 100 times, and each patient’s composite was averaged across repeats. For comparison, each patient’s NfL slope was estimated by regressing log_2_ NfL on time and sign-reversed, so that a positive correlation indicates that change tracks decline. The associations of the cross-fitted composite and of NfL slope with ALSFRS-R slope were compared using Pearson correlation, and their difference was tested with 2,000 paired bootstrap resamples. The contribution of the composite beyond NfL was assessed by comparing linear models of ALSFRS-R slope on baseline NfL and NfL slope with and without the composite using an F test. The comparison was then repeated after dividing patients into three groups of equal size by their baseline NfL level (low, intermediate, and high).

### Tracking across rates of decline

Patients were divided into thirds of equal size by their ALSFRS-R slope. Within each third, the composite was regressed on ALSFRS-R slope, so that patients were grouped on the predictor and not on the outcome, and the difference between the fastest and slowest thirds was tested by the interaction between ALSFRS-R slope and third. Robustness analyses and clinical comparisons between thirds are described in the Supplementary Methods.

### Replication in Target ALS

Differences between patients with ALS and controls were assessed in ‘limma’ with adjustment for sex and age, and for TMT set on mass spectrometry. Proteins were matched to Answer ALS by gene symbol. Concordance was assessed as the proportion changed in the same direction, tested against 50% with a two-sided binomial test, and by Pearson correlation of effect sizes. For progression, ALSFRS-R slopes (n = 133) and protein slopes (n = 50) were estimated using the same mixed-effects models. Concordance with the Answer ALS progression markers was assessed in the same way. A composite of the progression markers was then calculated with each marker’s direction fixed from Answer ALS, so that no marker was selected in Target ALS, and was related to ALSFRS-R slope by Pearson correlation with a one-sided permutation *p*-value. This was compared with 1,000 composites of the same number of randomly selected non-marker proteins.

### Statistical analyses

Bootstrap 95% CIs were calculated from 2,000 resamples. All analyses were done in R (v4.5.1) using ‘dplyr’, ‘tidyr’, ‘lme4’, ‘lmerTest’, ‘limma’, ‘DESeq2’, ‘edgeR’, ‘GenomicRanges’, ‘ChIPseeker’, ‘TxDb.Hsapiens.UCSC.hg38.knownGene’, ‘org.Hs.eg.db’, ‘AnnotationDbi’, and ‘concordR’.

## Data Availability

The Answer ALS data analyzed in this study, including clinical data, plasma SomaScan proteomics, serum NfL, and iMN RNA-sequencing, mass spectrometry, and ATAC-sequencing, are available to researchers on application through the Answer ALS Data Portal (https://dataportal.answerals.org; AALS-01184). The Target ALS Global Natural History Study clinical data and plasma mass spectrometry and Olink data are available on application through the Target ALS Data Engine (https://targetals.org/research/target-als-data-engine/). No new data or materials were generated in this study. All results supporting the findings are provided in the paper and the Supplementary Materials. The code and results for all analyses and figures are publicly available at https://github.com/Art83/als_long. The code for the simulations comparing the two-stage design with alternative methods is publicly available at https://github.com/Art83/als_sim.

https://targetals.org/research/target-als-data-engine/

## Acknowledgements

The authors are grateful to the Answer ALS Consortium, clinical cohort contributors, patients, donors, and families who helped to make this research possible. Data used in the preparation of this article were obtained from the ANSWER ALS Data Portal (AALS-01184). For up-to-date information on the study, visit https://dataportal.answerals.org. Data was also sourced from the Target ALS Global Natural History Study. More information on the study can be found at the Target ALS Data Engine: https://targetals.org/research/target-als-data-engine/.

## Author contributions

Conceptualization, Methodology, Software, Validation, Formal analyses: A.S.

Investigation, Resources, Data Curation, Writing – Original Draft, Writing – Review & Editing, Visualization, Project Administration, Funding Acquisition: C.A.F. and A.S.

