## Supplementary Methods and Figures for "Amyotrophic lateral sclerosis progression is tracked in plasma by systemic proteins independent of motor neurons"

**Supplementary Materials**

**Supplementary Methods**

**Sample timing and NfL processing**

The time of each plasma draw was taken from the plasma collection date or, where missing, from the ALSFRS-R assessment at the same visit, which was identical to the collection date for 97.3% of draws with both. Fifteen of 824 dated ALS draws were timed this way, and five draws with neither date were excluded. For serum NfL, samples recorded as quantity not sufficient or not tested (n = 3) were excluded, one value above the upper limit of the reportable range (>500 pg/mL) was set to 500 pg/mL, and the days-from-baseline value recorded with the immunoassay data was used for the 10 draws without a visit date.

**Comparison of longitudinal designs by simulation**

Simulation parameters were estimated from Answer ALS. ALSFRS-R variance components were taken from the mixed-effects model fitted to the 663 patients with two or more assessments, and protein variance components from the same model fitted to 100 randomly sampled aptamers. The number and timing of ALSFRS-R assessments and plasma draws were drawn from the distributions observed in the 229 patients with two or more draws. Patients who declined faster had fewer assessments (r = 0.23), and dropout was calibrated to reproduce this. Simulated ALSFRS-R scores were bounded between 0 and 48. Each simulated cohort comprised 229 patients and 1,000 proteins, of which five were true progression markers. For these, the coefficient of protein slope on ALSFRS-R slope was 0.015 or 0.025 log₂ RFU per ALSFRS-R point and the correlation of baseline level with ALSFRS-R slope was 0.15 or 0.25.

The two-stage design with BLUP slopes was compared with eight alternative methods. Three used per-patient ordinary least squares (OLS) slopes in place of BLUP slopes, either alone, with markers selected on the BLUP slopes, or with the coefficient divided by the reliability of the ALSFRS-R slope (regression calibration). A repeated-measures mixed-effects model related ALSFRS-R score to protein level at the same visit with participant-specific random intercepts. Four used the baseline sample only, and these were the correlation of protein level with ALSFRS-R score at the same visit, the correlation of protein level with the pre-baseline rate of decline (ΔFS), the difference in protein level between patients declining faster and slower than the median, and a mixed-effects model of ALSFRS-R with an interaction between protein level and time. Power was the proportion of true markers with an FDR below 0.05, and the observed FDR was the proportion of proteins with an FDR below 0.05 that were not true markers. All methods were compared in 100 simulated cohorts, and the methods other than the interaction model in 50 simulated cohorts at each of 2, 3, 4, 6, 8, and 12 visits per patient.

The methods were then applied to the 229 patients in Answer ALS. Two further methods were included in the cohort, the two-stage design with BLUP used for the ALSFRS-R slope only and the correlation of baseline protein level with the ALSFRS-R BLUP slope. Reproducibility was assessed by dividing patients at random into two halves 100 times and calculating the proportion of the 20 aptamers ranked highest in one half that were also among the 20 ranked highest in the other. The coefficient of protein slope on ALSFRS-R slope is attenuated when both slopes are BLUPs (by 58 to 71% in simulation), so the two-stage design was used to identify progression markers and associations are reported as correlations.

**Motor neuron expression map**

RNA-sequencing counts were variance stabilized using a blind transformation in ‘DESeq2’ (*1*). Protein detection was called from the mass spectrometry matrix released before batch correction and imputation, and ATAC-sequencing used the DiffBind consensus peak count matrix provided by Answer ALS. Aptamers were mapped from UniProt accession to gene symbol and Ensembl identifier using ‘org.Hs.eg.db’. A plasma protein was considered detected at the transcript level if its gene had five or more counts in at least 20% of iMN lines and a mean variance-stabilized expression above the 10^th^ percentile across all genes, and at the protein level if it was quantified in at least 50% of iMN lines. Promoter peaks were defined as ATAC-sequencing peaks within 2 kb of a transcription start site using ‘ChIPseeker’ against the UCSC hg38 knownGene annotation (*2*), and a promoter was considered accessible if any of its peaks had more than five counts in at least 20% of lines. Sensitivity was assessed at protein detection thresholds of 30%, 50%, 70%, and 90% of lines, and at transcript thresholds of 10%, 20%, 30%, and 50% of lines with and without the expression floor and against the ‘edgeR’ filterByExpr rule (*3, 4*).

**Motor neuron expression and progression tracking**

Baseline signal is measured on the same assay as the tracking correlation, so the adjusted model was repeated with the Human Protein Atlas (*5*) mass spectrometry-based blood concentration estimate (log10 pg/L) in place of SomaScan signal, which was available for 3,181 aptamers. Among aptamers detected at the transcript level, tracking was compared between expressed and transcript-only plasma proteins using a Wilcoxon rank sum test and a linear model with the same adjustment. Enrichment of map classes was assessed using unique UniProt accessions, with the distribution across all five classes compared using Fisher’s exact test with a Monte Carlo *p*-value (100,000 replicates). The overlap between differential plasma proteins and progression markers, and its dependence on the direction of the difference between patients and controls, was assessed using Fisher’s exact test.

**Route to plasma and tissue distribution**

Each protein was assigned a route from its UniProt subcellular localization as secreted, membrane, intracellular, or unknown, with membrane proteins taken to reach plasma by shedding and intracellular proteins by release from damaged cells (*6*). The proportion of intracellular proteins relative to secreted and membrane proteins was compared with the panel using Fisher’s exact test. Protein presence was assessed across 59 human tissues by combining immunohistochemistry from the Human Protein Atlas (*7*), tissue mass spectrometry from the GTEx proteome (*8*), and PaxDb (*9*), and a protein was considered detected in a tissue if any source detected it. For each tissue, a presence weight was calculated as the immunohistochemistry level (high 3, medium 2, low 1, none 0) divided by 3, plus 1 each for detection in GTEx and PaxDb, and Shannon entropy of the scaled weights was normalized by the log_2_ of the number of tissues covered. Normalized entropy was compared between progression markers and the panel using a Wilcoxon rank sum test. The panel for these analyses comprised the 6,295 unique gene symbols on the SomaScan panel. Spinal cord is not represented in these atlases and could not be tested.

**NfL reliability and regression to the mean**

The reliability of the BLUP slopes for NfL and ALSFRS-R was estimated for each model as the mean across patients of one minus the ratio of the conditional variance of each patient’s slope to the between-patient slope variance. The observed correlation between NfL slope and ALSFRS-R slope was divided by the square root of the product of the two reliabilities (*10*). Baseline models were fitted with and without adjustment for baseline severity, represented by each patient’s random intercept from the ALSFRS-R model. To test for regression to the mean, NfL slopes were re-estimated from follow-up draws only in patients with three or more draws, with each patient kept in the baseline NfL group assigned from their first draw. The difference in correlation between the low and high groups was then tested using Fisher’s z transformation.

**Robustness and clinical characteristics of fast progressors**

For each marker, the regression coefficient within the fastest and slowest thirds was divided by its coefficient across all patients. Follow-up length was added as a modifier, and the analysis was repeated in patients followed for at least one year. Protein and ALSFRS-R slopes were re-estimated after excluding samples taken within six or 12 months of death, and the comparison was repeated with patients divided into halves, quartiles, and quintiles. The pre-baseline rate of decline (ΔFS) was calculated as 48 minus the baseline ALSFRS-R score, divided by months since symptom onset (*11*). ALSFRS-R subdomains were calculated as the sums of the bulbar (items 1 to 3), fine motor (4 to 6), gross motor (7 to 9), and respiratory (R1 to R3) items. Vital capacity was the highest percent predicted forced vital capacity at each visit, or slow vital capacity where forced vital capacity was not recorded. Slopes of each subdomain, vital capacity, and BMI were estimated using the same mixed-effects model. A C9orf72 repeat expansion was defined as a repeat length of 30 or more on whole-genome sequencing or a clinically reported expansion. Characteristics were compared using Wilcoxon rank sum and Fisher’s exact tests, with FDR controlled across characteristics. Within each third, the cross-fitted composite was related to the slope of each subdomain, vital capacity, and BMI by Pearson correlation, and within the fastest third this was compared between onset sites by an interaction term. The ability of baseline NfL and ΔFS to identify patients in the fastest third was assessed by the area under the receiver operating characteristic curve (AUC).

**Target ALS processing**

Normalized TMT reporter ion ratios were log_2_-transformed, and technical replicates for the same visit were averaged over the channels in which each protein was quantified. Proteins quantified in fewer than 50% of samples were removed. TMT set was included as a covariate as TMT ratios are relative to other samples in the same set. Olink values were available at a single visit and were not used for progression. Where a gene was measured by more than one aptamer in Answer ALS, the aptamer with the smallest *p*-value was used. Protein slopes were tested only for proteins measured in at least 15 patients. For the composite, progression markers measured by a single-target aptamer were matched to TMT proteins by UniProt accession, and patients with slopes for fewer than half of the markers were excluded. The 95% CI was calculated from 2,000 bootstrap resamples and the permutation p-value from 2,000 permutations of ALSFRS-R slope.

**Supplementary Figures**

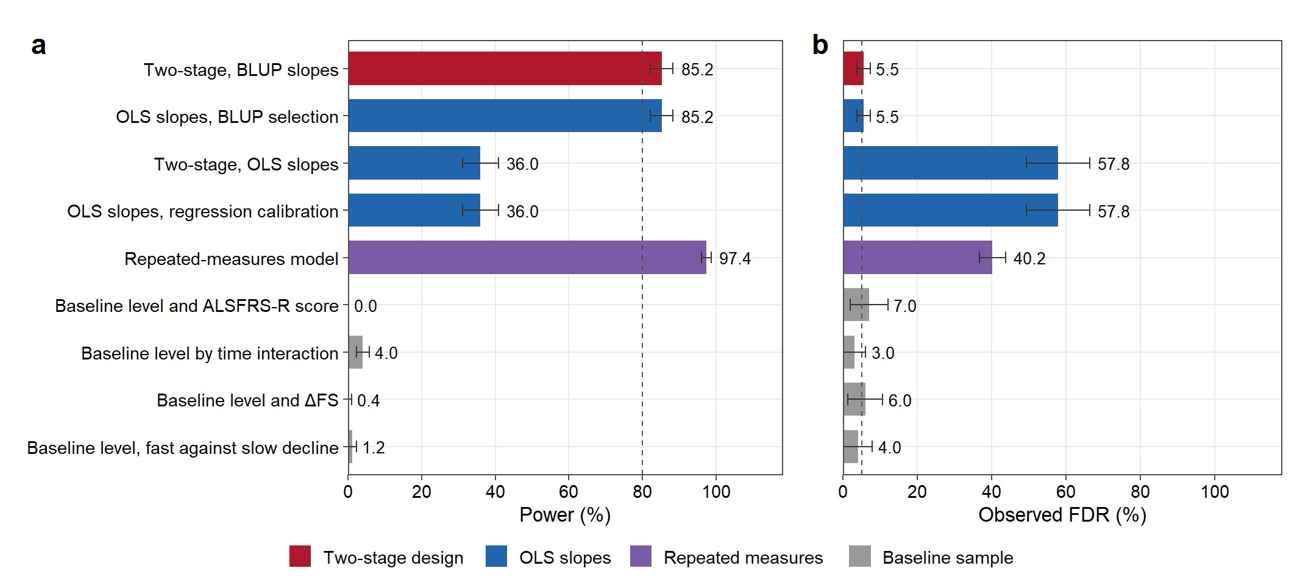

**Figure S1. The two-stage design detects true progression markers while controlling the false discovery rate in simulated cohorts.** (**A**) Power and (**B**) observed FDR of each method in cohorts of 229 patients simulated from Answer ALS, each with five true progression markers among 1,000 proteins. Bars show the mean across 100 simulated cohorts. The dashed lines mark 80% power and an FDR of 5%.

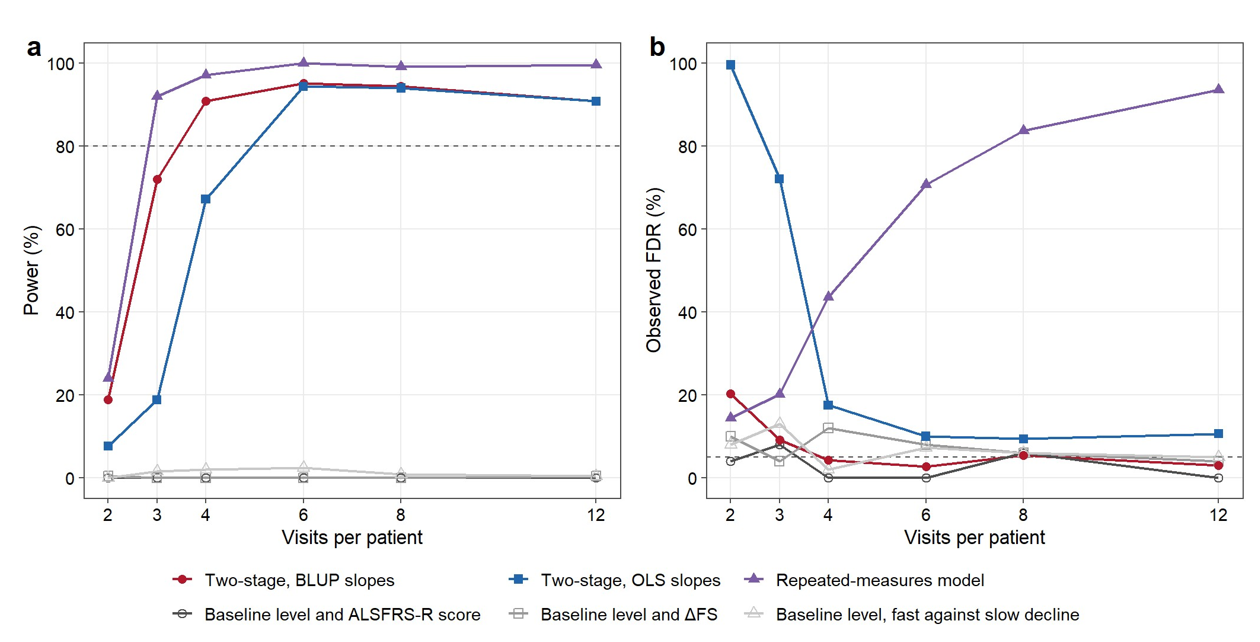

**Figure S2. Power and false discovery rate depend on the number of visits per patient.** (**A**) Power and (**B**) observed FDR of each method in cohorts simulated with 2, 3, 4, 6, 8, or 12 visits per patient. Points show the mean across 50 simulated cohorts. The dashed lines mark 80% power and an FDR of 5%.

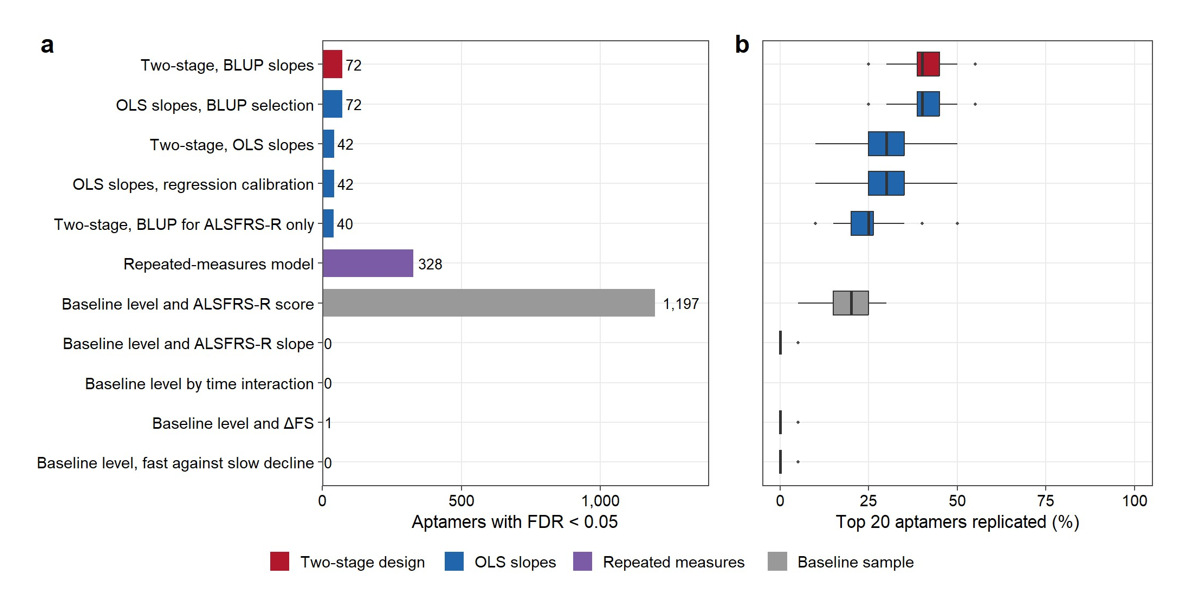

**Figure S3. Progression markers from the two-stage design replicate between random halves of the cohort more often than those from alternative methods.** (**A**) Number of aptamers with an FDR below 0.05 for each method in the 229 patients in Answer ALS. (**B**) Proportion of the 20 aptamers ranked highest by each method in one random half of the cohort that were also among the 20 ranked highest in the other half, across 100 random divisions. Boxes show the median and interquartile range.

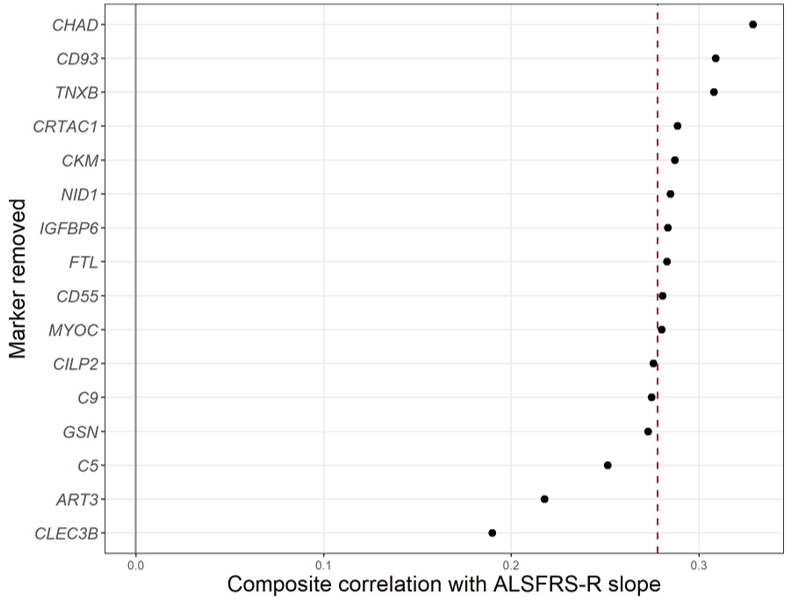

**Figure S4. The association of the composite with decline in Target ALS is robust to the removal of any single progression marker.** Correlation between change in the composite of progression markers and ALSFRS-R slope in Target ALS after removal of each of the 16 markers in turn. The dashed line marks the correlation of the composite with all 16 markers included.

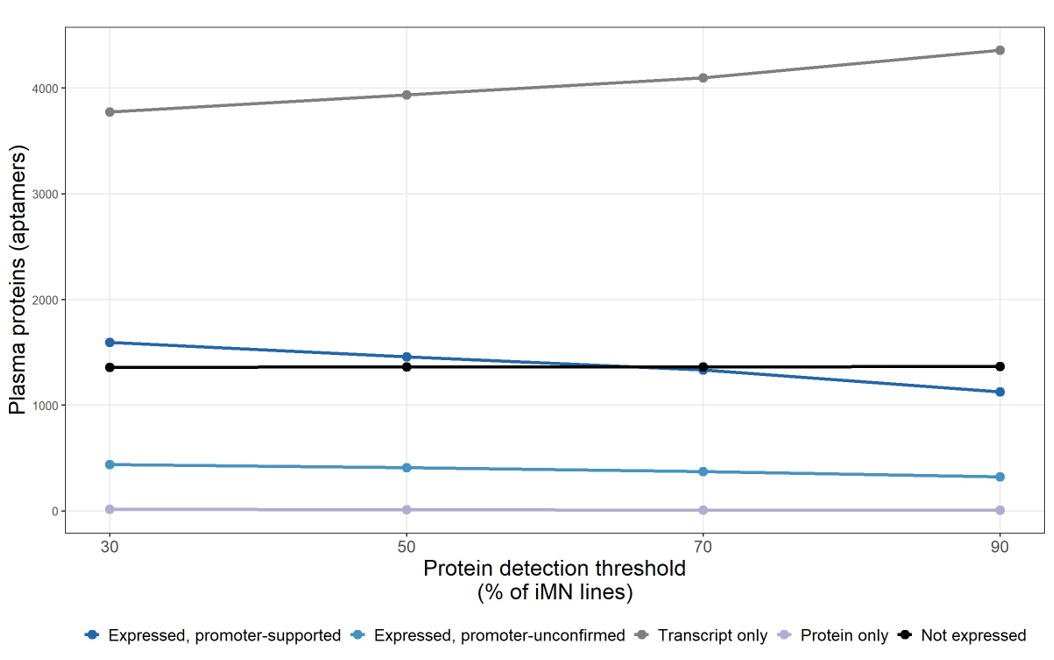

**Figure S5. The classes of the motor neuron expression map are stable across protein detection thresholds.** Number of plasma proteins in each class of the map when a plasma protein was considered detected at the protein level if it was quantified in at least 30%, 50%, 70%, or 90% of iMN lines. Counts are given at the aptamer level. The 50% threshold was used in all other analyses.

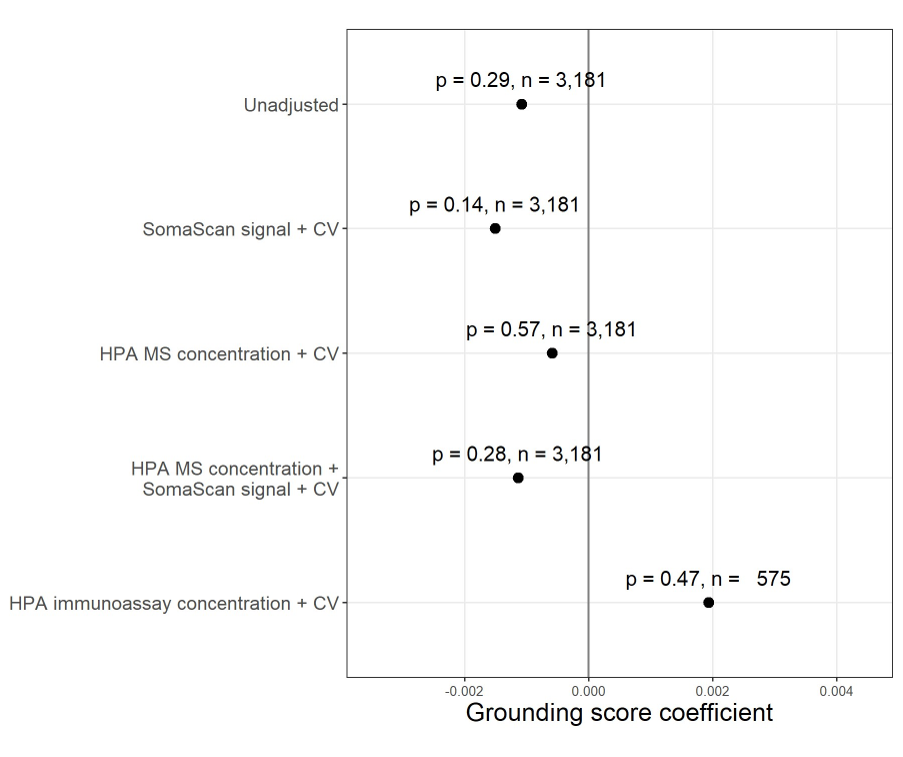

**Figure S6. Grounding score is not associated with progression tracking after adjustment for external blood concentration.** Coefficient of grounding score in linear models of progression tracking, shown without adjustment and with adjustment for SomaScan baseline signal, for the Human Protein Atlas (HPA) mass spectrometry (MS)-based blood concentration estimate, for both, and for the HPA immunoassay-based blood concentration estimate. All adjusted models included the coefficient of variation (CV) of baseline signal. The *p*-value and number of aptamers are given for each model.

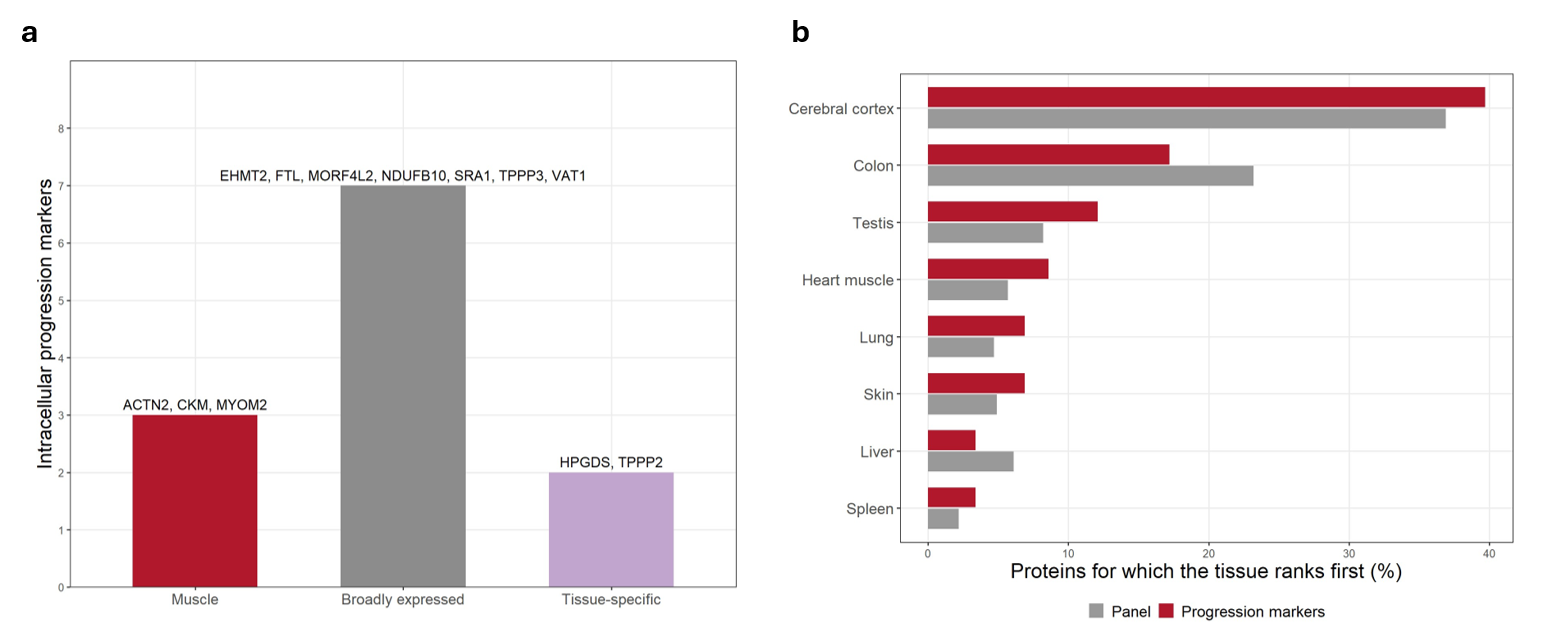
**Figure S7. Intracellular progression markers are not neuronal, and cerebral cortex ranks first for a similar proportion of progression markers and of the panel.** (**A**) Number of the 12 intracellular progression markers that were muscle proteins, broadly expressed proteins, or tissue-specific proteins. The proteins in each group are named above each bar. (**B**) Percentage of the progression markers (red) and of the panel (gray) for which each tissue ranked first in concordR.

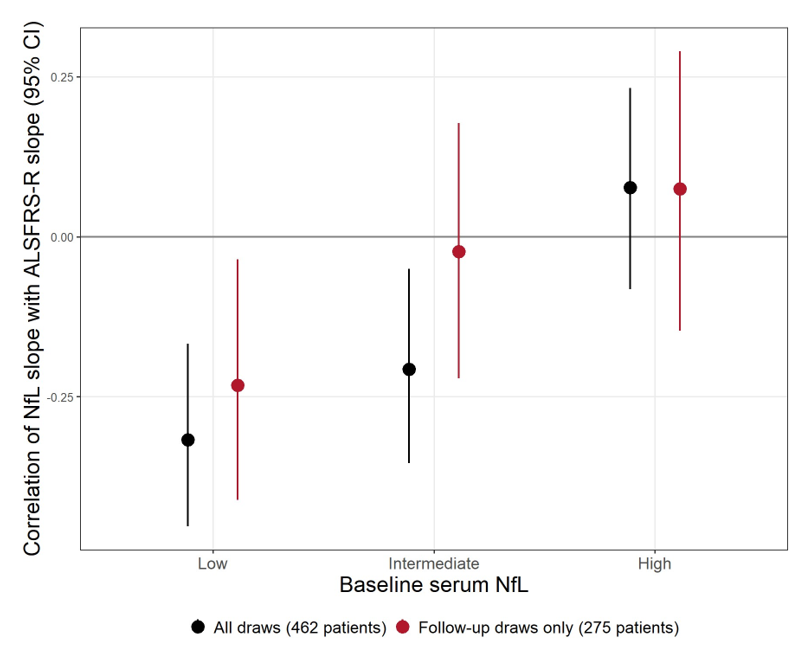

**Figure S8. Change in NfL tracks decline in patients with low baseline NfL when slopes are estimated from follow-up draws alone.** Correlation between NfL slope and ALSFRS-R slope in patients with low, intermediate, and high baseline serum NfL. Slopes were estimated from all draws in patients with two or more draws (black, n = 462) and from follow-up draws only in patients with three or more draws (red, n = 275), with each patient kept in the group assigned from their baseline draw. Points and bars show the correlation and its 95% CI.

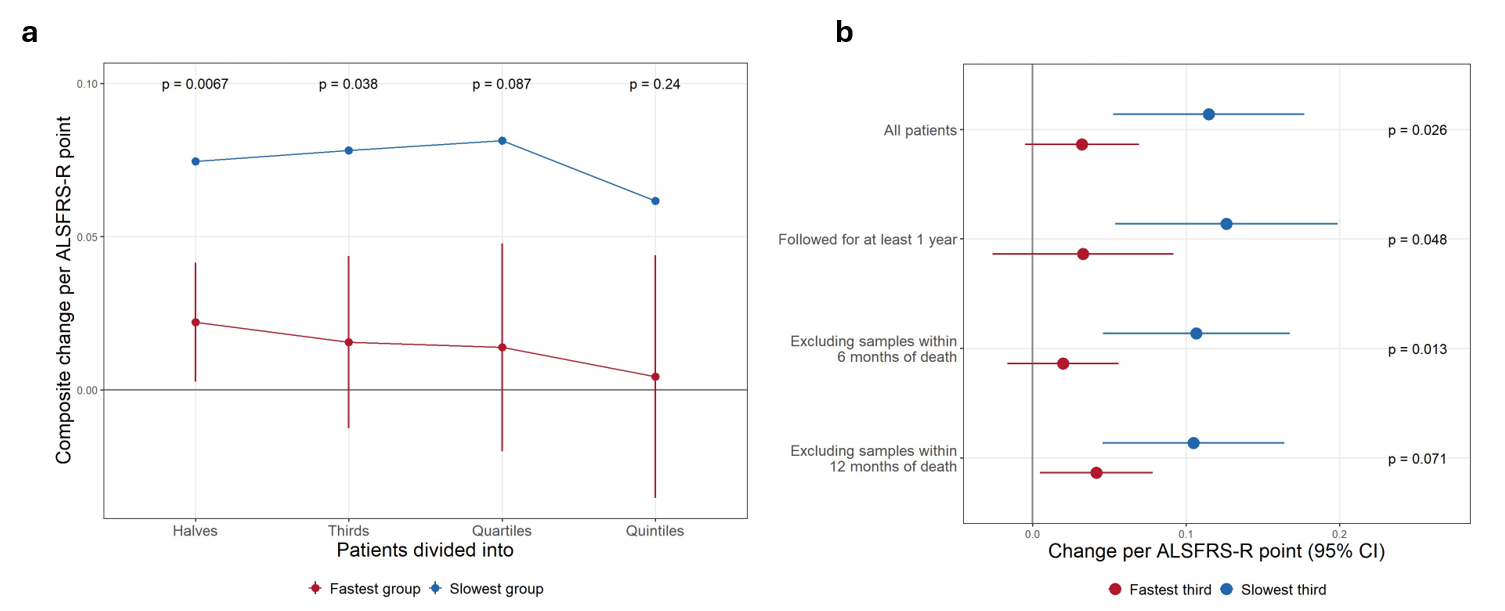

**Figure S9. The loss of tracking in the fastest progressing patients is robust to how patients are grouped, to follow-up length, and to samples taken close to death.** (**A**) Change in the cross-fitted composite per ALSFRS-R point per year in the fastest (red) and slowest (blue) groups when patients were divided into halves, thirds, quartiles, and quintiles by their rate of decline. Bars show the 95% CI for the fastest group, and the p-value for the difference between the two groups is given above each division. (**B**) Change in the composite per ALSFRS-R point per year in the fastest (red) and slowest (blue) thirds in all patients, in patients followed for at least one year, and after excluding protein and ALSFRS-R samples taken within six or 12 months of death. Points and bars show the coefficient and its 95% CI, and *p*-values are for the difference between the fastest and slowest thirds.
